# Contact patterns and HPV-genotype interactions yield heterogeneous HPV-vaccine impacts depending on sexual behaviours: an individual-based model

**DOI:** 10.1101/2021.02.23.21252238

**Authors:** Mélanie Bonneault, Chiara Poletto, Maxime Flauder, Didier Guillemot, Elisabeth Delarocque-Astagneau, Anne CM Thiébaut, Lulla Opatowski

**Author notes:** **Author for correspondence:** Mélanie Bonneault, Institut Pasteur, Epidemiology and Modelling of Antibiotic Evasion Unit, 25, rue du Docteur-Roux, 75475 Paris Cedex 15, France. These authors contributed equally to the research (Anne CM Thiébaut, Lulla Opatowski).

## Abstract

Human papillomaviruses are common sexually transmitted infections, caused by a large diversity of genotypes. In the context of vaccination against a subgroup of genotypes, better understanding the role of genotype interactions and human sexual behavior on genotype ecology is essential. Herein, we present an individual-based model that integrates realistic heterosexual partnership behaviors and simulates interactions between vaccine and non-vaccine genotypes. Genotype interactions were considered, assuming a previous vaccine-genotype infection shortened (competition) or extended (synergy) the duration of a secondary non-vaccine-genotype infection. Sexual behavior determined papillomavirus acquisition and transmission: only 19.5% of active individuals with 0–1 partner during the year, but >80% of those with ≥2 partners, were infected before vaccine introduction. Genotype interactions, despite being silent during the pre-vaccination era, markedly impacted genotype ecology after vaccination started, with a significant increase/decrease of non-vaccine prevalence for competitive/synergistic interactions. These changes were more pronounced in individuals with ≤3 partners (up to 30% of prevalence modification assuming 65% vaccine coverage) but barely visible for individuals with >3 partners (at most 0.30%). Results suggest that considering genotype interactions, in conjunction with heterogeneous sexual behaviors, is essential to anticipate the impact of existing and future anti-papillomavirus vaccines targeting a subgroup of genotypes.

## Introduction

Human papillomavirus (HPV) genital tract infections are among the most common sexually transmitted infections, especially in younger people (de Sanjosé et al., 2007; Markowitz et al., 2013). HPVs are characterized by a high diversity of genotypes, a few of which have been associated with cancers (cervical, vaginal, vulval, penile and anal) (Walboomers et al., 1999). Prevention has recently relied on vaccination with bivalent, quadrivalent and, more recently, nonavalent (Serrano et al., 2012) vaccines that target the subtypes carrying the highest carcinogenic risks.

Following the introduction of anti-HPV vaccines, HPV-infection prevalences with targeted genotypes have declined in several countries (Drolet et al., 2019; Tota et al., 2020). However, quantifying the vaccines’ impacts on global HPV prevalence remains difficult. In particular, an increased prevalence of genotypes not included in the vaccine, known as genotype replacement, is a potential risk. Therefore, when vaccinating against a subgroup of genotypes, prediction of vaccine impact requires better understanding of whether genotypes interact and, if so, how.

In fact, simultaneous, within-host co-infection by two genotypes can affect each virus genotype’s load, cell-infection ability and/or infection duration (McLaughlin-Drubin and Meyers, 2004; Murall et al., 2014; Xi et al., 2009). On the one hand, some evidence indicates that HPV genotypes may compete (Biryukov and Meyers, 2018; McLaughlin-Drubin and Meyers, 2004; Xi et al., 2009); on the other, multiple infections were commonly observed before vaccine introduction (Chaturvedi et al., 2011; Mejlhede et al., 2010; Mendez et al., 2005; Spinillo et al., 2009). The mechanisms leading to the co-existence of widely diverse HPV genotypes in populations have not yet been definitively elucidated. Some researchers argue co-existence is consistent with genotypes acting independently of each other (Chaturvedi et al., 2011; Vaccarella et al., 2013), while others suggest that the observed clustering of distinct HPV genotypes is due to between-genotype interactions (Mejlhede et al., 2010; Merikukka et al., 2011; Spinillo et al., 2009).

Previous HPV-transmission models formalizing interactions between HPV genotypes have not yet been able to reproduce simultaneously the complex aspects of HPV biology and individual behaviors. So far, models accounting for genotype interactions have been based on homogenous-mixing assumptions (Elbasha and Galvani, 2005; Pons-Salort et al., 2013; Poolman et al., 2008), although partnership may vary greatly across individuals and ages. These heterogeneities, in addition to affecting infection risk and HPV prevalence (Shiboski and Padian, 1996), could also markedly impact genotype co-infection, co-circulation and interaction dynamics at the population level. Indeed, theoretical study results suggested that the contact network affects the population ecology of pathogen strains (Eames and Keeling, 2006; Pinotti et al., 2019). Gray *et al*. consistently observed HPV-genotype-replacement differences between individuals with high-risk and low-risk behaviors (Gray et al., 2019). Therefore, an accurate accounting of sexual behavior might be essential to correctly interpret HPV observations and provide more accurate projections of the epidemiological and ecological consequences of vaccination.

Optimally, only individual-based models (IBMs) enable reproduction of the heterogeneity of individual behaviors and simulation of their effects at population levels (Auchincloss and Diez Roux, 2008). A few IBMs have been developed for HPV (Johnson et al., 2018; Matthijsse et al., 2015; Olsen and Jepsen, 2010; Van de Velde et al., 2012, 2010) but none has considered HPV-genotype interactions. Herein, we report how the interplay between genotype interactions and host behaviors determine genotype prevalence, co-infection and vaccine impact using an HPV-transmission IBM based on a realistic heterosexual network of individuals and accounting for the most prevalent HPV vaccine (V) and non-vaccine (NV) genotypes.

## Materials and methods

### Global overview

We developed a stochastic HPV-transmission IBM accounting for contact heterogeneity according to sex, sexual activity and age. We considered the 14 most prevalent genotypes, including two V (HPV-16 and -18), and 12 NV genotypes (HPV-31, -33, -35, -39, -45, -51, -52, -56, -58, -59, -66 and -68) and modelled their transmissions individually and simultaneously, exploring a range of competitive and synergistic genotype interactions. The model assumed natural acquired immunity and included vaccination implementation, assuming the vaccine provided full immunity against V-genotype-HPV infections.

### Population and individual characteristics

Individuals of both sexes enter the population at 15 years and leave it at 30 years. Individuals who exit the population are directly replaced by new 15-year-old individuals, keeping the population stable over time. The population is divided equally by sex and age per year. Each individual is explicitly modelled for each week and characterized by his/her age, sex, partnership status (three categories: in a relationship, available for a new partnership, or neither) and infection status for each genotype (four categories: susceptible, infected, naturally immunized or vaccinated in case of a V genotype).

### Modelling partnership

We modelled heterosexual partnership behaviors as a stochastic process according to the steps summarized in figure 1 and detailed in supplementary material S1.

**Figure 1.**
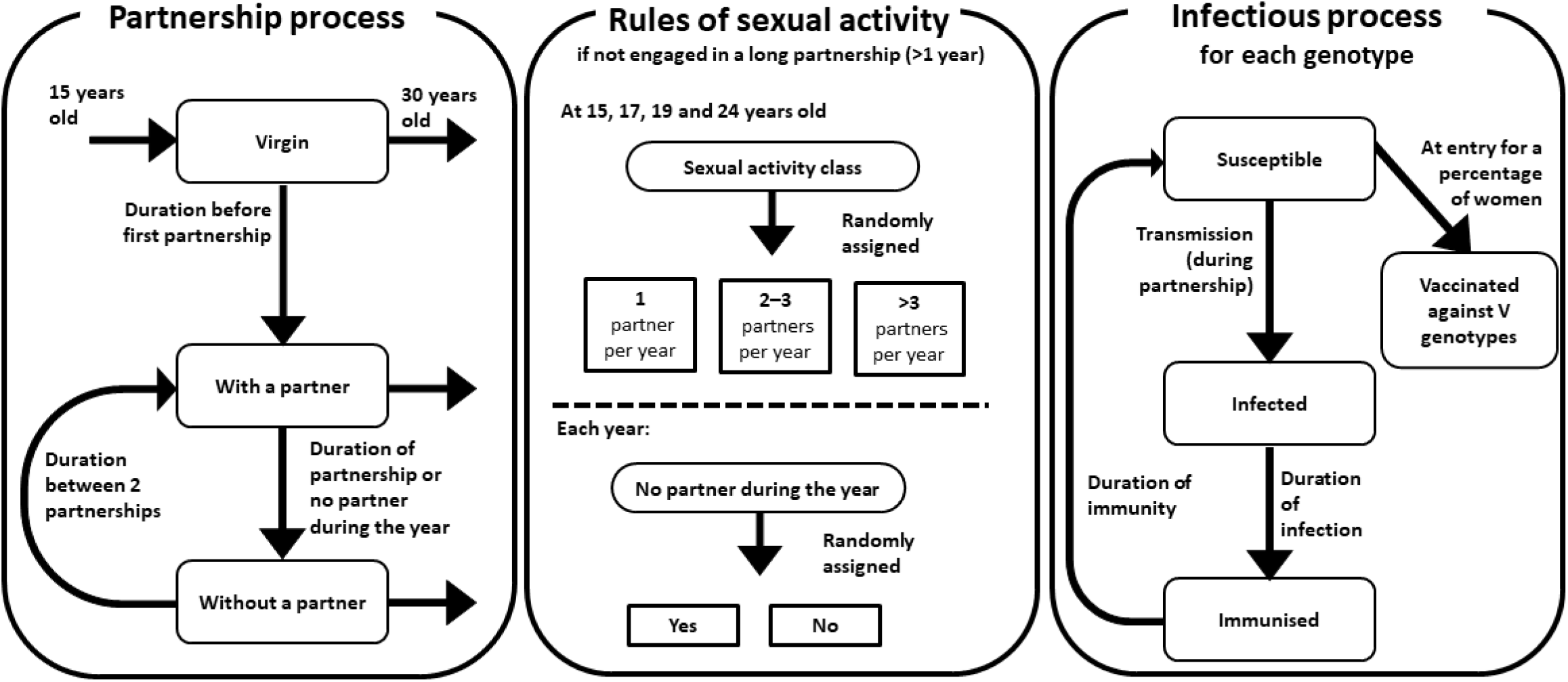
Schematic representation of partnership and HPV infectious processes. The partnership process is guided by the rules of sexual activity defining active and inactive periods.

Model parameters describing the partnership were estimated to reproduce the reported contact patterns from the *Contexte de la Sexualité en France* (CSF) survey (Bajos and Bozon, 2008). The calibrated sexual behavior parameters are summarized in supplementary material S2.2. Combinations of parameter values were simulated. After reaching prevalence equilibrium, we extracted the cumulative number of sexual partners from age 15 years to that at the time of data extraction for each individual. Because women’s and men’s behaviors could not be reproduced simultaneously, we first selected the combinations consistent with women’s data, then chose the best combination among them with respect to men’s data.

### Modelling HPV-genotype transmission and infection

The transmission of any of the 14 genotypes can occur when an infected individual is in partnership with an individual not infected with that specific type (figure 1). Transmission-probability parameters are defined for V and NV genotypes, respectively *β*_*V*_ and *β*_*NV*,_ for two contacts/week. If the virus is transmitted, acquisition occurs. If an individual is infected—regardless of the genotype—his/her infection duration is sampled from an exponential distribution of mean 52 weeks, in accordance with the literature (Trottier et al., 2008). After infection clearance, the individual is assumed to have acquired natural immunity, conferring total protection against the same genotype for a duration defined by an exponential distribution. At the end of that period, the individual again becomes fully susceptible to that genotype. The transmission-probability parameters, *β*_*V*_ and *β*_*NV*,_ and mean immunity duration were calibrated to reproduce prevalence before vaccine introduction (Markowitz et al., 2013), first assuming genotypes to be independent with respect to transmission and infection (neutral interaction scenario, details in supplementary material table S4, in S2.3).

With those hypotheses, the 14 viruses spread simultaneously among partners in the population. Figure 2 illustrates transmission over a simulated network.

**Figure 2.**
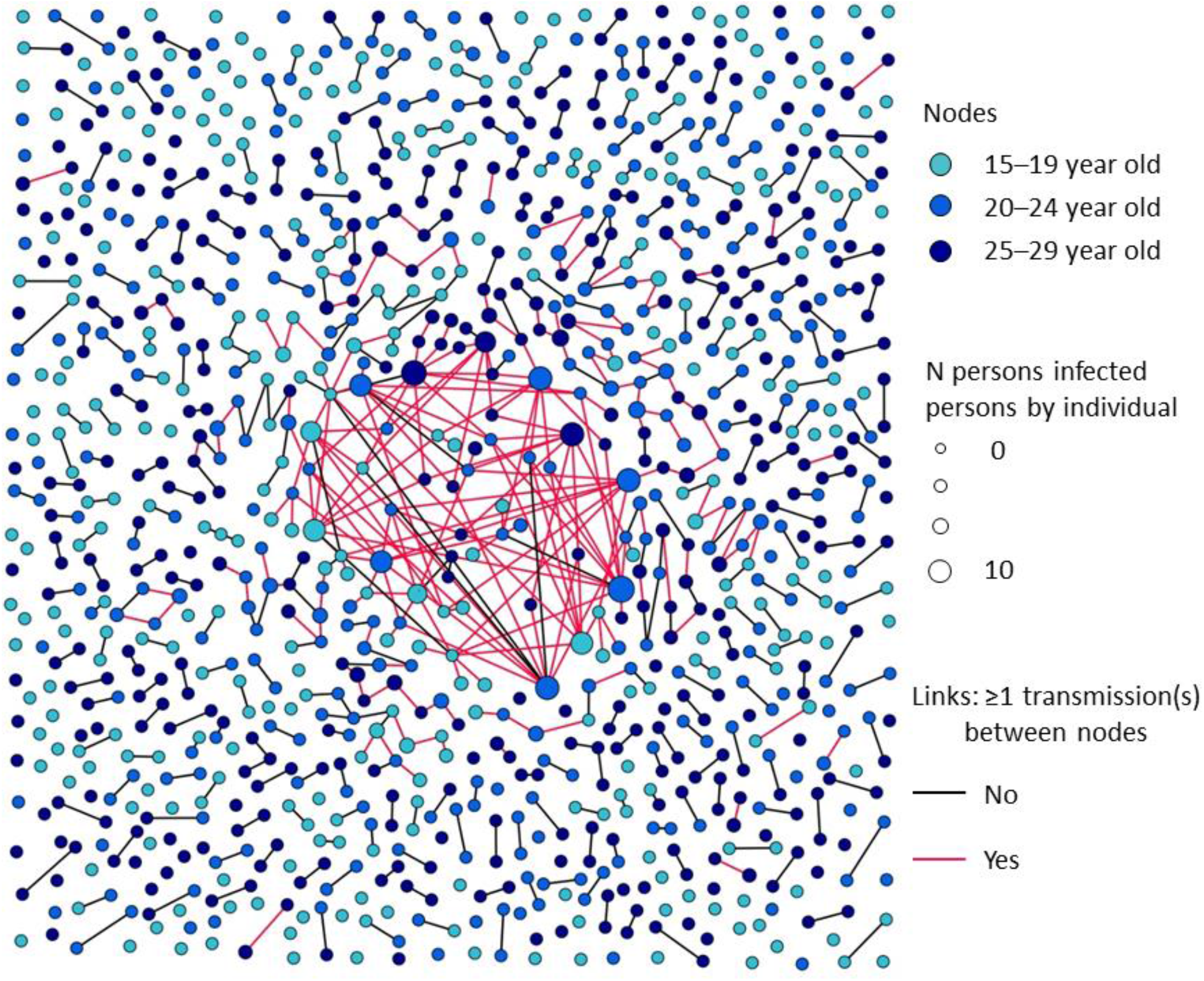
Partnership and HPV-transmission network throughout 1 year with a simulated population composed of 1058 individuals and 565 connections. Node colors correspond to age categories. Node diameters correspond to the number of persons infected by the individual over the 1-year study period after reaching prevalence equilibrium. If two individuals were in relationship during the year, a linking line is drawn between them, colored in red if ≥1 transmission(s) occurred between them, otherwise black.

### Between-genotype interaction

We focused only on V–NV-type interactions. When an individual is co-infected with two HPV genotypes, we assumed that their co-infection history could be affected and considered the following interaction mechanism: when an individual is already infected with a V genotype, the duration of a consecutive NV-genotype infection is multiplied by *α*, the strength of interaction. We considered five interaction scenarios: the neutral interaction scenario (*α* = 1) (see above), two scenarios with synergistic interaction in the range of values that could be satisfactorily calibrated (*α* = 1.25 and 1.5) and two symmetrical competitive interaction scenarios (*α* = 0.8 and 0.67). We also explored alternative hypotheses of interaction (changing infection acquisition instead of duration, bilateral between V- and NV-genotype groups or universal between any genotypes instead of unilateral V/NV interaction) at one competition (0.8) and synergy level (1.25) as sensitivity analyses in supplementary material S6.

### Vaccine introduction

We focused on the immunization schedule against high-risk genotypes that had prevailed for more than a decade in most countries (ECDC, 2020). We assumed that immunization conferred full protection (100% efficacy) against HPV-16 and -18 genotypes for at least 15 years. We introduced vaccination with varying coverage after prevalence equilibrium was reached and ran the model for 50 additional years. We also examined the impact of targeting <15-year-old women with the highest infection and transmission risks as an additional sensitivity analysis (results in supplementary material S4.6).

### Parameter calibration

We used the least-squares distance minimization to measure the adequacy between simulated results and real data. All parameters, calibrated or taken from the literature, are defined in table 1. Ranges of tested values and estimated values are detailed in supplementary material S2 and table S4 for each calibrated parameter.

**Table 1.**
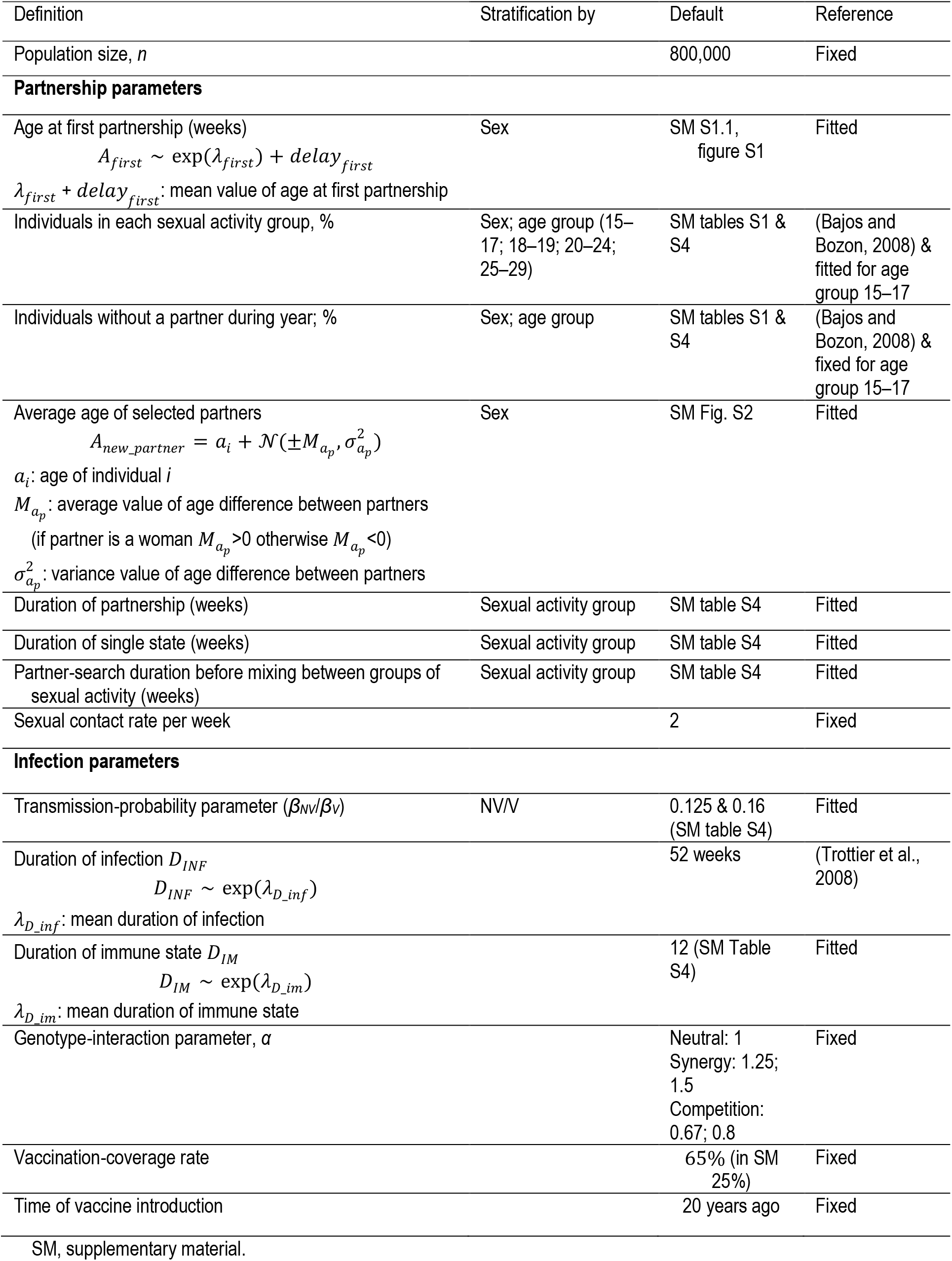
Model parameters

### Analysis of results

To characterize V- and NV-genotype distributions, we considered infection, co-infection, acquisition and transmission according to sexual activity groups (detailed in supplementary material S3.2). First, we compared HPV-infection spread through the contact network for varying genotype-interaction strengths with that under the neutral interaction scenario before vaccine introduction. Second, we examined how hypothetical genotype-interaction scenarios would affect HPV-genotype ecology after vaccine introduction in comparison with the neutral interaction scenario (detailed methods in supplementary material S3.3).

## Results

### Model simulations reproduced observed partner and HPV-prevalence distributions by age

Compared to CSF results, distributions of total numbers of partners were reproduced with limited variability (figure 3a), minor overestimation of the percentage of women with >10 partners and underestimation of the percentage of men with >3 partners (figure S3).

**Figure 3.**
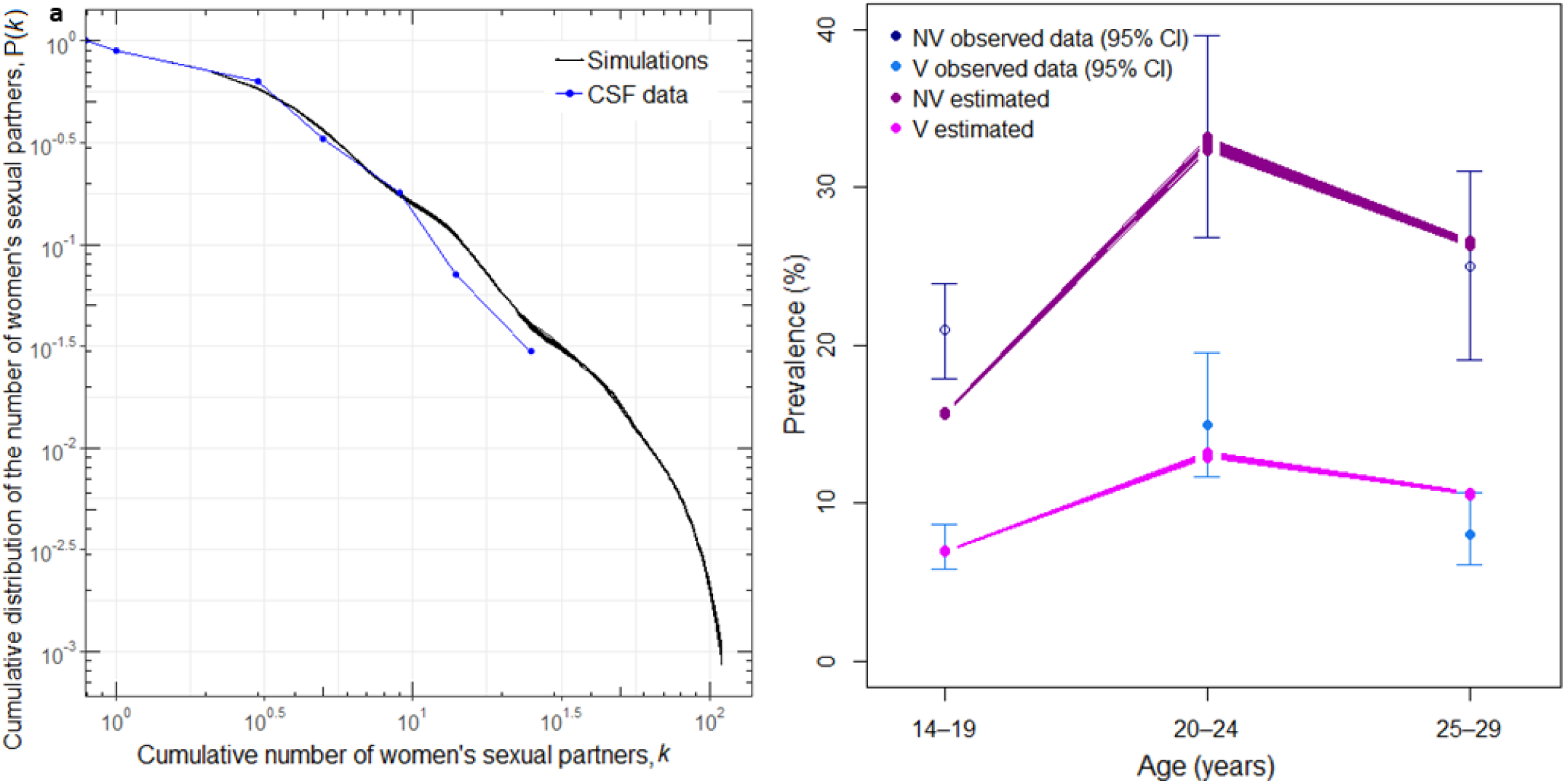
Calibration of partnerships and HPV-transmission parameters. (a) Cumulative distribution of the number of sexual partners for women 18–30 years old. (b) NV- and V-genotype prevalences with 95% confidence interval (CI) reported in *(Markowitz et al*., *2013)* (circles) and results of 10 simulations, each obtained as the average value over 10 years after reaching prevalence equilibrium (connecting lines).

Under the neutral interaction scenario, the model accurately reproduced realistic distributions of V- and NV-genotype prevalences by age category, with the characteristic bell-shaped curves and prevalence peaks between 20 and 24 years (figure 3b). The transmission-probability parameter calibrated for V genotypes was higher than that of NV genotypes (*β*_*V*_ = 0.16 and *β*_*NV*_ = 0.125, respectively, with 12 weeks of calibrated immunity duration). In simulations, the average percentage of ever-infected individuals increased sharply with age, reaching 71.4% at 29 years.

### Infections and co-infections did not spread homogeneously throughout the network under the neutral interaction scenario

Individuals infected with a single or multiple genotype(s) were distributed among the population as a function of the number of partners during the past year (figure 4a). After reaching prevalence equilibrium, 33.3% of the active population were infected (27.7% of the whole population) and 20.3% were co-infected (61.5% of the infected). Among the infected population (figure 4b), 45.2% of infected individuals had 0–1 partner during the past year (15.5% prevalence for this group), 43.2% had 2–3 partners (81.7%) and 11.6% had >3 partners (99.5%). The co-infection percentage increased with annual sexual activity, with only 8.2% of active individuals with 0–1 partner being co-infected, as opposed to >60% of individuals with ≥2 partners. Co-infections with multiple V genotypes but no NV genotype were extremely rare (0.35% of infected individuals). Indeed, 33.1% of the infected population were V–NV-genotype co-infected.

**Figure 4.**
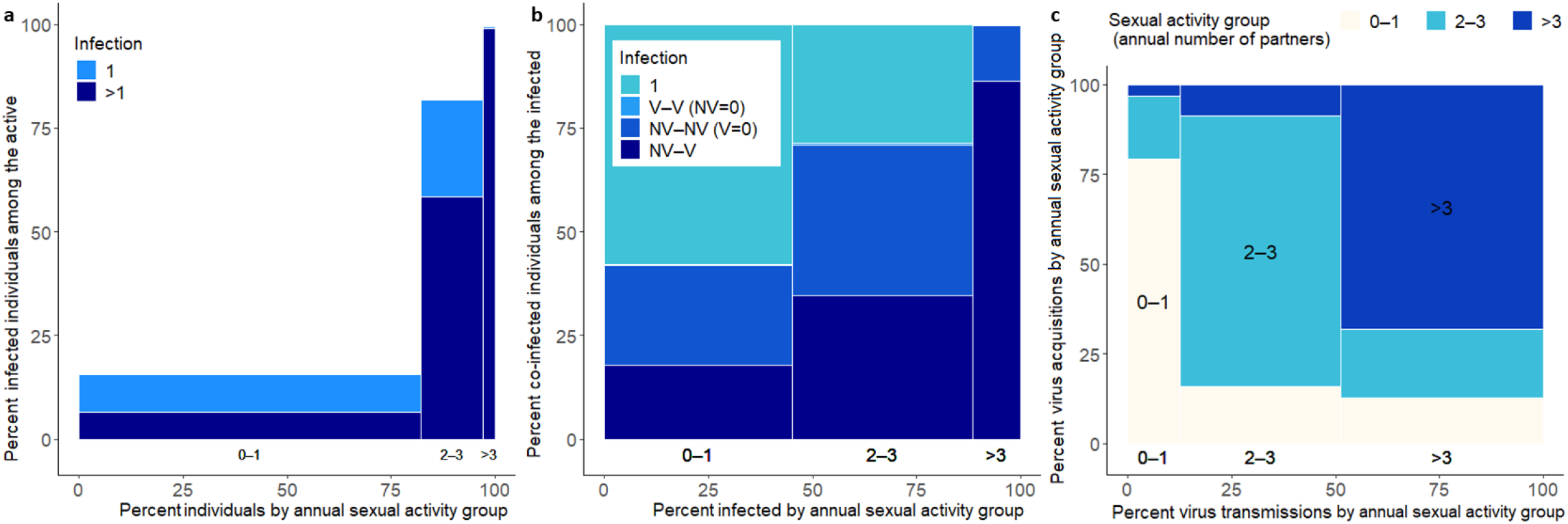
NV- and V-genotype infections, co-infections and transmissions according to the annual number of partners. (a) Mean percentages of individuals with single and multiple infections among the active population over 20 simulations according to their number of partners during the past year at prevalence equilibrium *t* = 640 weeks. (b) Mean percentages of individuals infected with a single NV or V genotype, with the two V types but no NV type, with >1 distinct NV types but no V type, and with V and NV types among the infected population over 20 simulations according to their annual number of partners at prevalence equilibrium time. (c) Percentages of virus transmissions according to sexual activity group of the transmitter during the year. For each transmission group (x-axis), the sexual activity group distribution of the acquirers is represented on the y-axis.

As shown in figure 4c, HPVs were mostly transmitted by individuals with ≥2 partners (41.2% by individuals with 2–3 partners and 47.5% by individuals with >3 partners in the year); 95.5% of individuals with >3 partners and 48.3% of individuals with 2–3 partners transmitted at least once during the year. Moreover, transmission mostly occurred within the sexual activity group (comparison with partnership matrix in supplementary material S4.2).

### Alternative genotype-interaction scenarios did not modify pre-vaccine infection and co-infection patterns

Before vaccination was introduced, all simulations were fitted to HPV-genotype prevalences within the range of assessed competitive, neutral and synergistic interactions (figure S5).The strength of interaction did not markedly affect co-infection patterns either (figure S8a, b and c). We further verified that this lack of influence did not simply reflect differences in transmission probability *β*_*NV*,_ resulting from calibrating the model for varying interaction strengths (supplementary material S5.2).

### V and NV prevalences were impacted differently according to sexual activity group post-vaccine introduction

After introducing vaccination, hypotheses regarding genotype-interaction impacted NV-prevalence trends (figures 5 and S6 for a uniform 65% vaccine coverage among women; results for 25% coverage are given in supplementary material S4.5). In all genotype-interaction scenarios, V-prevalence range decreased (difference relative to pre-vaccine era) by 83.7% to 85.9% for individuals with 0–1 and 2–3 partners, and only 51.3% to 53.2% for individuals with >3 partners (figures S6 and 5a). Conversely, NV-genotype prevalences changed markedly after vaccine introduction for all interaction strengths compared to the neutral scenario. Competitive or synergistic interaction scenarios, respectively, led to higher or lower NV prevalence compared to the neutral scenario (figures S6, and S9a and d). As expected, the stronger the interaction strength (the more it deviates from 1), the greater the NV-prevalence percentage difference (for additional information, see the supplementary material on sensitivity analysis S5.4). The NV-prevalence effect was stronger in individuals with 0–1 partner (median 25.9% for *α* = 0.67 and –32.0% for *α* = 1.5) than in individuals with 2–3 partners (median 14.2% for *α* = 0.67 and –23.5% for *α* = 1.5) (figure 5b). For individuals with >3 partners during the year, prevalence did not change appreciably (median 0.08% for *α* = 0.67 and –0.25% for *α* = 1.5).

**Figure 5.**
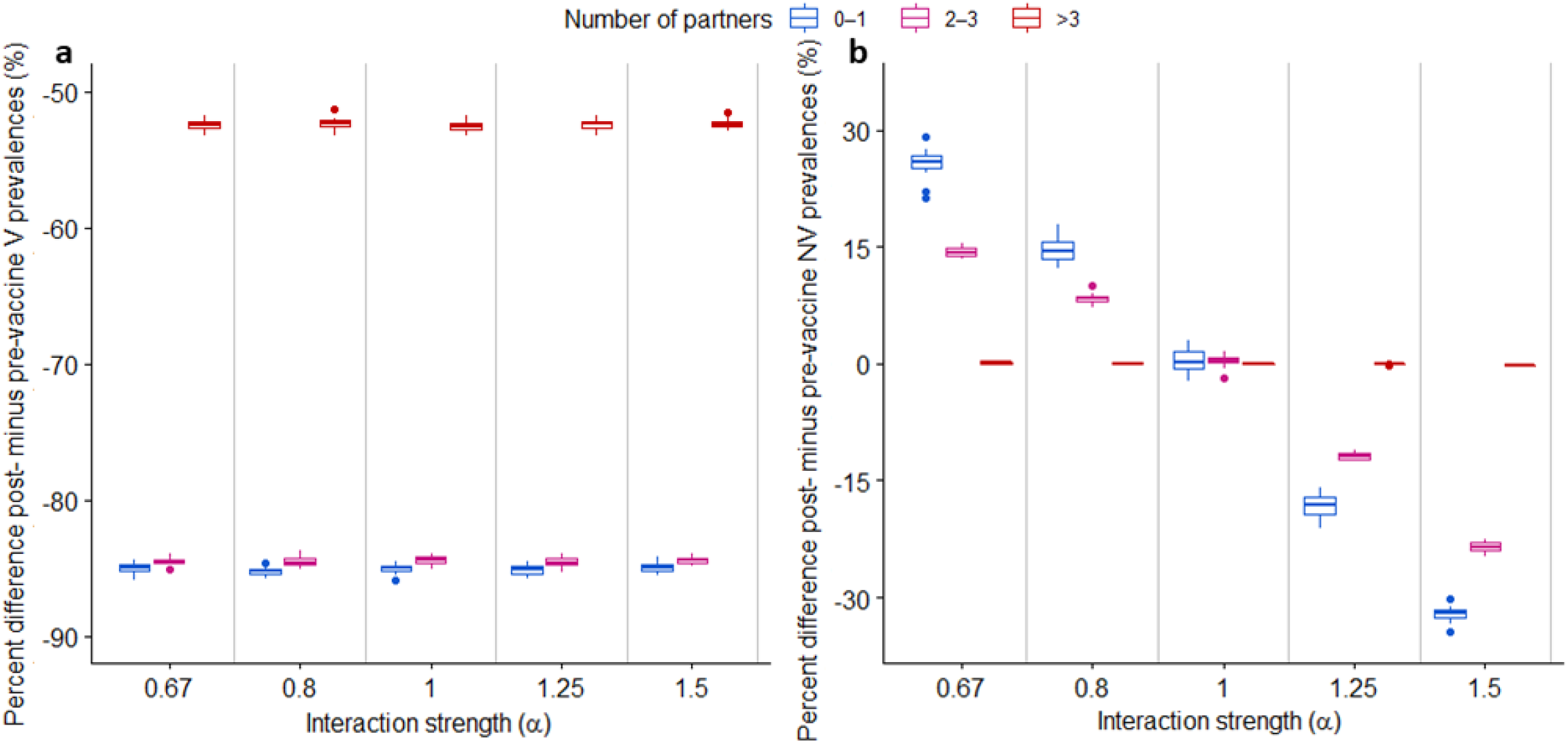
Percentage differences between post- and pre-vaccine prevalences of (a) V and (b) NV genotypes under 65% vaccine coverage by sexual activity group and interaction scenarios. Vaccine was introduced after reaching prevalence equilibrium. Average prevalences were assessed at three distinct times each before and after vaccine introduction. Boxplots show the variability over 20 simulations for each interaction scenario (bold horizontal line inside the box is the median, lower and upper box limits are the 1st and 3rd quartiles, antennae correspond to 1.5 times the interquartile range and circles mark the values outside this range).

Sensitivity analyses showed that targeting high-risk sexual activity groups instead of vaccinating all women <15 years old led to greater reduction of V-type prevalence for individuals with >3 partners but did not modify NV-prevalence-variation patterns across sexual activity groups (supplementary material S4.6). Furthermore, among the other hypotheses tested for genotype interactions (supplementary material S6), it appeared that only interactions by HPV-genotype groups could be detected within the considered range of interaction strengths. Moreover, when notable NV-prevalence differences were observed, compared to the neutral interaction scenario, they appeared only among the less active individuals.

## Discussion & conclusion

We found that heterogeneous contact behaviors and natural immunity acquired after infection were essential to reproduce the bell-shaped curve of HPV-infection prevalence by age category. Calibrated interaction scenarios revealed no notable differences in terms of infection and co-infection patterns before vaccine introduction. However, thereafter, interactions mattered: competitive interactions led to significantly increased NV prevalence, while the opposite was seen for synergistic interactions. Of interest, our results suggested that the interaction effect on prevalence was stronger for less active individuals.

While it is expected that the introduction of the nonavalent vaccine will again modify the prevalence equilibrium, solid comprehension of the potential impacts of the initial bi- and quadrivalent vaccines is required to improve understanding of changing HPV ecology. Our IBM allowed us specifically to examine the impact of vaccination scenarios of V-, NV-or V–NV genotype interactions. Two former studies based on compartmental models analyzed the impact of genotype interactions on acquisition or clearance, without explicitly modelling heterogeneous sexual behaviors as IBMs do (Elbasha and Galvani, 2005; Pons-Salort et al., 2013). The results of both showed that, when considering between-genotype interactions, NV-genotype prevalences could be modified by vaccine introduction.

Based on our simulations for the prevaccine era, 71.4% of individuals were infected with at least one HPV genotype by 29 years of age. This simulation finding agrees with previous reports of 80% infected at least once during their lifetime (Santé publique France, 2019), acknowledging that a limited number of new infections can be acquired after 30 years of age and that only 14 high-risk HPV among 37 detectable sexually transmitted genotypes were modelled here. Furthermore, co-infections were frequent in our results, representing about two-thirds of HPV-positive women for all interaction strengths tested. That outcome, although at the upper limit of the interval, is consistent with 20%–70% co-infections among all infections described in observational studies (Chaturvedi et al., 2011; Mejlhede et al., 2010; Mendez et al., 2005; Spinillo et al., 2009). Pertinently, herein, all co-infections were assumed to be detected and counted, unlike epidemiological studies in which, depending on the techniques used, detection of multiple infections may be more-or-less sensitive (Qu et al., 1997).

Concerning transmission probabilities under the neutral interaction scenario, our estimated *β*_*NV*_- and *β*_*V*_-transmission probabilities were 0.125/ and 0.16/week, respectively. Considering two sexual acts per week on average, our results are of the same order of magnitude as values from other IBMs, range 0.048–0.95 per sex act (Johnson et al., 2018; Matthijsse et al., 2015; Olsen and Jepsen, 2010; Van de Velde et al., 2012, 2010). Nevertheless, our estimates strongly depend on infection duration and the definition of immunity. Notably, assuming a probability <1 to acquire type-specific, natural, lifelong immunity yielded higher transmission probabilities, range 0.5–0.95 (Johnson et al., 2018; Van de Velde et al., 2012, 2010). In addition, either with acquired immunity during a limited period, as assumed herein, or without immunity, modelling showed lower transmission probabilities, range 0.048–0.3 (Matthijsse et al., 2015; Olsen and Jepsen, 2010).

Pertinently, the pre-vaccine situation could be explained by totally distinct interaction hypotheses. Moreover, vaccine introduction more strongly impacted NV prevalence in less active individuals, whereas any prevalence modification would be expected to be seen in high-risk groups (Gray et al., 2019; Tota et al., 2013). Considering transmission throughout the network, the acquisition risk for any individual depends on two factors: the virus-circulation level in the population (defined as the transmission rate, and durations of infection and immunity), and the number of partners. As those numbers remain unchanged on average, the vaccination impact depends on virus-transmission potential before vaccination. That potential was substantially higher for the >3-partner group than the low-or intermediate-activity groups.

Our results obtained with this methodology should also be interpreted in light of the following limitations. First, aggregated data were used, impeding a precise description of individual trajectories throughout their sexual lifetimes, such as switching between sexual activity groups, and durations of partnership and between two partnerships. As previously stressed in other studies, our model was unable to perfectly reproduce the distributions of the numbers of partners for both sexes. Indeed, CSF data included more men with high numbers of partners than women (Bajos and Bozon, 2008). Differential sex-dependent reporting bias was observed previously (Fenton et al., 2001; Mitchell et al., 2019) and may, in part, be at the origin of the discrepancy between model and data. To overcome that difficulty, we focused on women’s data to fit our model and estimate parameters, while trying to conform to men’s data in terms of cumulative partner numbers. The resulting distribution of total numbers of partners was typical of a partnership network with a power law distribution (Liljeros et al., 2001; Schneeberger et al., 2004).

Second, because no data are available on HPV-infection prevalence by age and V genotype for the pre-vaccine era in France, we used distributions reported for the US, whose current HPV epidemiology is similar (Bruni et al., 2010; de Sanjosé et al., 2007). NV-prevalence simulations appeared to be underestimated for 15–19-year-olds, compared with US data (Markowitz et al., 2013). Unfortunately, partnership and infection data are scarce for this age category, making calibration difficult. We think that underestimation for 15–19-year-olds had little impact on our results for all ages combined. The average prevalence of 27.7% for the whole 15–30-year-old population obtained in our simulations before vaccination is comparable to those reported in epidemiological studies in France, range 25.1%–28.5% for 15–79-year-old women with normal vaginal smear cytology (Dalstein et al., 2003; Monsonego et al., 2005; Riethmuller et al., 1999).

Third, to keep our model relatively simple, only two transmission-probability parameters were estimated for all genotypes, despite studies having reported that HPV-16 and -18 prevalences may differ from one another and from those of other HR genotypes (Markowitz et al., 2013). Hence, our estimated transmission-probability parameter *β*_*V*_ can be considered an average probability for HPV-16 and -18, and that of *β*_*NV*_ an average probability for the 12 high-risk NV types. Similarly, because epidemiological study results have not yet demonstrated clear between-genotype differences for infection duration (Ramanakumar et al., 2016; Trottier et al., 2008), a unique parameter was defined for all genotypes.

Fourth, while much is still unknown about immunity following HPV infection (Gravitt and Winer, 2017), we chose to define immunity as total protection against reinfection to the same genotype during a fixed duration, identical for all genotypes, in accordance with findings suggesting some degree of protection against reinfection (Ho et al., 2002; Safaeian et al., 2010). Previous HPV modelling considered either complete lifelong immunity, partial or waning immunity, acquired immunity during a defined period, or protection increasing with the number of past infections (Franceschi and Baussano, 2014). Matthijsse *et al*. showed that assuming natural immunity in their model was necessary to reproduce the age-specific HPV-infection patterns and that assuming full genotype-specific immunity yielded the best calibration results (Matthijsse et al., 2015). We made the same observation herein, after testing various immunity hypotheses (data not shown).

Finally, while interaction mechanisms between HPV genotypes also remain unknown, we assumed that genotype interaction would affect the infection duration of a second acquired virus. Some authors suggested that simultaneous within-host co-infection with two genotypes could impact each virus-genotype load and thus the successful infection of cells by each (Biryukov and Meyers, 2018; McLaughlin-Drubin and Meyers, 2004; Xi et al., 2009). We explored alternative interaction hypotheses in sensitivity analyses and obtained similar results for NV genotypes, when genotype interaction impacted acquisition risk instead of infection duration.

To conclude, our results confirmed that HPV infections reached not only the most sexually active but also less active individuals and unexpectedly revealed that the impact of vaccine introduction on genotype ecology could be more detectable in less active individuals. These analyses suggest that focusing on more sexually active individuals to address questions related to vaccination impact might potentially lower the ability to detect HPV-ecology variations, and highlight that public health information and prevention efforts should target all sexually active individuals, not only the population at-risk for sexually transmitted infections. Moreover, our results showed that genotype-interaction-associated hypotheses were all consistent with the reported pre-vaccine prevalences. In the post-vaccine era, better understanding those interactions and sexual contacts is key to anticipating the long-term impact of anti-HPV vaccines with respect to the prevention of cervical and other cancers.

## Data Availability

All of the code necessary to simulate the model and generate the figures will be made available on request

## Authors’ contributions

MB, CP, MF, EDA, DG, ACMT and LO conceptualized the project. AT and LO supervised the project. MB, ACMT and LO designed the model. MB and MF developed the model. MB, CP, ACMT and LO performed the network and statistical analyses and validated them. All authors approved the latest version of this article

## Acknowledgements

We thank Margarita Pons-Salort for helpful discussions and ideas provided early during the project. We are also grateful to Janet Jacobson for editorial assistance.

## Appendix A. Supplementary data

Supplementary data associated with this article can be found, in the online version.

