## Appendix A. Supplementary data for "Contact patterns and HPV-genotype interactions yield heterogeneous HPV-vaccine impacts depending on sexual behaviours: an individual-based model"

#### S1. Complementary information regarding partnership modelling

The partnership process is divided into several steps, as detailed below.

##### S1.1 Age at first partnership

At entry into the population, individuals are assigned an age at first partnership $A_{first}$ drawn from an exponential distribution according to the individual's sex.

$$A_{first} \sim \exp(\lambda_{first}) + delay_{first}$$

Mean $\lambda_{first}$ and intercept $delay_{first}$ (time interval to first partnership) were fitted by sex so that percentages of sexually active individuals by age fit to the *Contexte de la Sexualité en France* (CSF) data (Bajos and Bozon, 2008). Best fits to the CSF data were obtained for means of 3.8 and 4.0 years and time intervals of 13.0 and 13.5 years for men and women, respectively (figure S1).

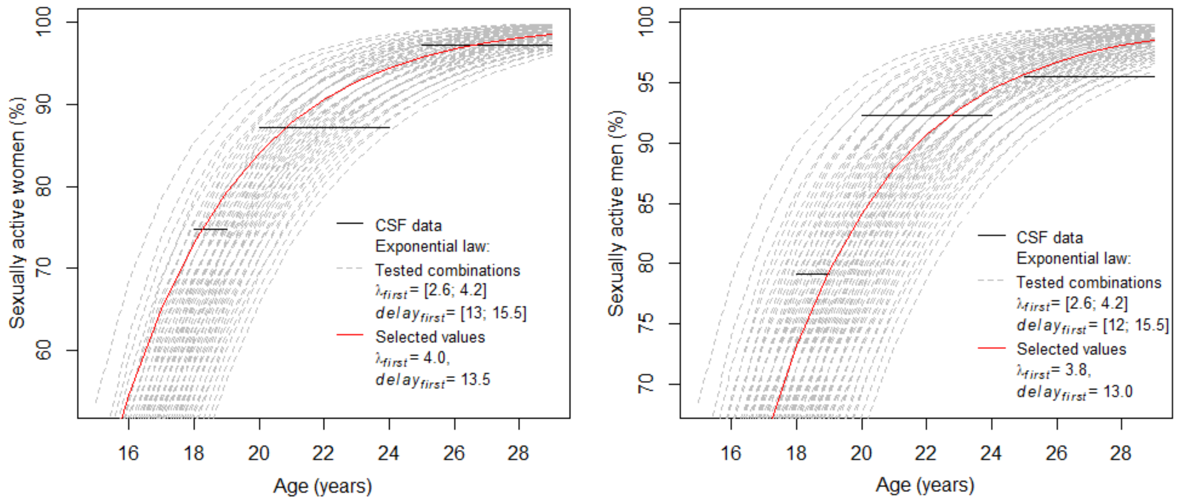

**Figure S1. Percentages of sexually active women (left) and men (right) according to age**: CSF data in *(Bajos and Bozon, 2008)*, simulated values over the defined interval are shown in grey and selected values in the best fit are shown in red. $\lambda_{first}$ represents the average values of the exponential law and the time interval to first partnership $delay_{first}$.

##### S1.2 Sexual activity, classes and rules

For sexually active individuals, a sexual activity class (three categories: 1 (c1), 2–3 (c2) or >3 partners per year (c3)) is randomly assigned according to age (table S1). These classes determine the duration of active and inactive periods to obtain heterogeneous behaviours. Distributions of individuals across sexual activity classes according to sex and age were fixed using CSF data, except for age group 15–17 years, whose percentages were fitted by sex to reproduce cumulative percentages of sexually active individuals by age and sex in the CSF survey (due to the lack of data).

At ages 17, 19 and 24 years, a portion of individuals, not engaged in long partnerships (>1 year), were randomly selected to be assigned a sexual activity class according to the percentage of individuals in each sexual activity class by age category. This procedure enabled the percentage of individual in each group according to age category to be respected. A maximum percentage of individuals who change groups $MC_c$ is defined and fitted by sex to reproduce the cumulative numbers of partners per age group and sex (Bajos and Bozon, 2008).

Additionally, each calendar year, a defined percentage of individuals was randomly selected to have no partner during that year, regardless of activity class.

**Table S1.** Distributions of individuals by sex, age and sexual activity class according to CSF survey (Bajos and Bozon, 2008)

| Sex | Number of partners per year | | | |
|---|---|---|---|---|
| Age group (years) | 1 (c1) | 2–3 (c2) | ≥4 (c3) | 0* |
| Women | | | | |
| 18–19 | 68.99 | 24.72 | 6.29 | 3.63 |
| 20–24 | 75.84 | 21.56 | 2.60 | 7.24 |
| 25–29 | 85.75 | 11.07 | 3.18 | 5.59 |
| Men | | | | |
| 18–19 | 60.42 | 27.59 | 11.99 | 14.38 |
| 20–24 | 61.85 | 28.15 | 10.00 | 10.89 |
| 25–29 | 76.30 | 17.23 | 6.47 | 6.69 |

*Individuals without a partner are drawn from the whole population each year.

##### S1.3 Finding a partner

When an individual is ready to form a new partnership, a partner of the opposite sex is sought by sampling among all active individuals of the same sexual activity class, within a span of possible ages, taking into account age differences between the two partners. This span, defined as the average age $A_{new\_partner}$ of the new partner, is sampled in a normal law:

$$A_{new\_partner} = a_i + \mathcal{N}(\pm M_{a_p}, \sigma_{a_p}^2)$$

where $a_i$ is the age of individual *i*, $M_{a_p}$ is the mean of the normal law (positive for a male partner, negative for a woman) and $\sigma_{a_p}^2$ is the variance fitted to reproduce the distributions

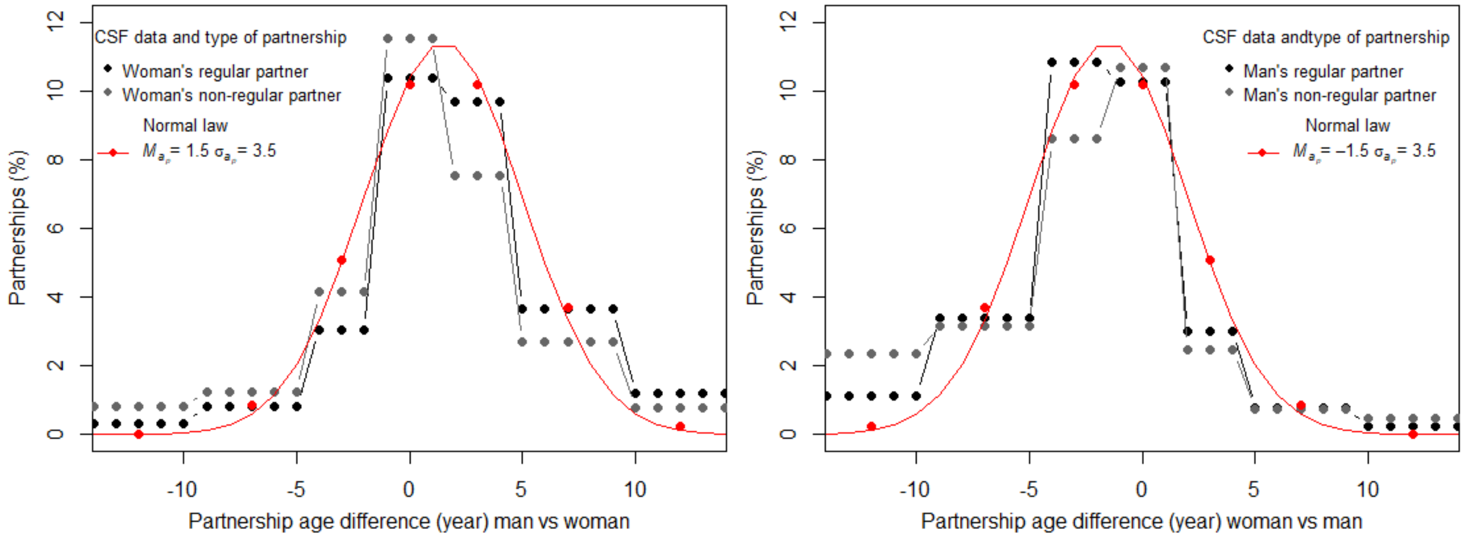

**Figure S2. Percentages of partnerships according to age difference between the man and woman (left) and the woman and man (right).** CSF data are represented in black for regular partner (spouse or main partner) and in grey for non-regular (new or casual partner). The calibration result is in red.

of the between-partner age differences from CSF data (Bajos and Bozon, 2008). The obtained mean values were +1.5 years for a woman, –1.5 years for a man and 3.5 for variance (figure S2). The possible age range was derived from this average age span that is a more-or-less fixed number of weeks. If individuals do not find an appropriate partner within their sexual activity class after a calibrated search time, then mixing between sexual activity groups is allowed.

##### S1.4 Durations of partnerships and between partnerships

When a partnership is formed, its duration is defined according to the sexual activity class of the most active of the two partners. If both partners belong to the same sexual activity class, it is the individual who finds his/her partner who determines the partnership's duration. No other concurrent partnership was allowed.

At the end of their partnership, individuals stay single for a given length of time, defined according to their respective sexual activity class.

For individuals with a theoretically unique partner during the year, durations are determined in two steps. First, a duration is randomly selected according to the percentage of individuals by sex with partnership duration <1, 1–4, 5–9 or ≥10 years in the CSF data (Bajos and Bozon, 2008) (table S2). Then a value within that time span was selected from a uniform random law.

The duration between two partnerships is calculated so that they cannot have another partner during the same year:

$$D_S = 52 - \left(day(t) - next_{year}\right) + U[1:52]$$

**Table S2.** Distribution of partnership durations by sex and age category, for individuals with 1 partner per year, CSF survey (Bajos and Bozon, 2008)

| Sex | Partnership duration (years) | | | |
|---|---|---|---|---|
| Age group (years) | ≤1 | 1–4 | 5–9 | ≥10 |
| Men | | | | |
| 18–19 | 36.32 | 26.78 | 20.79 | 16.11 |
| 20–24 | 30.15 | 26.30 | 24.64 | 18.91 |
| 25–34 | 6.76 | 9.74 | 43.26 | 40.23 |
| Women | | | | |
| 18–19 | 24.62 | 40.45 | 20.00 | 14.93 |
| 20–24 | 17.35 | 27.01 | 26.95 | 21.54 |
| 25–34 | 3.76 | 10.14 | 44.07 | 42.02 |

For individuals with several partners during the year, partnership and between-partnership durations were selected from gamma laws calibrated to sex. To obtain heterogeneous behaviours among individuals with >3 partners, durations of partnerships and between-2-partnerships were selected at random only once a year. The sampled values for each individual then apply to any partnership newly created within the year.

When individuals turn 30, they exit the population. If they are partnered at that time, their partner stays in the relationship during the duration defined at pair formation.

#### S2. Simulation and parameter-calibration values

##### S2.1 Computer simulations

The model was simulated for a population of 800,000 individuals with a 1-week interval.

The model was developed in C++, simulations were run on the computational and storage services (TARS cluster) provided by the IT Department at Institut Pasteur, Paris. Statistical analyses and graphics were computed using R (version 3.5.2).

##### S2.2 Partnership-parameter calibration

Partnership parameters, which were optimised to reproduce cumulative numbers of partners by age category, were the durations of partnership and between two partnerships, percentages of individuals staying in the same class, percentages of 15–17-year olds in each group, and persistent partner-search duration before mixing between sexual activity classes (table S4). For each combination of parameters, the distributions by categories of the number of sexual partners since age 15 years (0, 1, 2–3, 4–5, 6–9, 10–14, 15+ partners), age category (18–19, 20–24 and 25–29 years) and sex obtained by simulations were compared with CSF survey data (table S3) (Bajos and Bozon, 2008).

**Table S3.** Distributions of numbers of lifetime partners by sex and age at the time of the CSF survey [1]

| Sex | Number of lifetime partners | | | | | | | |
|---|---|---|---|---|---|---|---|---|
| Age (years) | 0 | 1 | 2–3 | 4–5 | 6–9 | 10–14 | ≥15 | Unknown |
| Men | | | | | | | | |
| 18–19 | 20.9 | 24.4 | 23.2 | 11.4 | 8.9 | 4.8 | 6.4 | 0 |
| 20–24 | 7.7 | 14.8 | 25.3 | 15.6 | 15.6 | 9.2 | 11.1 | 0.8 |
| 25–29 | 4.4 | 13.2 | 14.9 | 16.5 | 14.0 | 15.4 | 19.8 | 1.8 |
| Women | | | | | | | | |
| 18–19 | 25.1 | 25.5 | 31.4 | 8.7 | 5.4 | 3.2 | 0.5 | 0.3 |
| 20–24 | 12.8 | 28.1 | 31.5 | 14.8 | 7.8 | 2.7 | 2.3 | 0 |
| 25–29 | 2.7 | 24.0 | 28.6 | 17.8 | 15.8 | 5.9 | 4.6 | 0.6 |

##### S2.3 Calibration of infection and interaction parameters

The transmission-probability parameters, $\beta_V$ and $\beta_{NV}$, and the mean duration of immunity were calibrated to reproduce prevalence before vaccine introduction in a US study (Markowitz et al., 2013) under the initial assumption that genotypes are independent with respect to transmission and infection (neutral interaction scenario, table S4). We considered V- and NV-genotype prevalences among women by age category (14–19, 20–24 and 25–29 years), defined as the ratio of the number of women infected with at least one V- or NV-group genotype over the total number of women in the same age category. To smooth fluctuations, we averaged the V and NV prevalences by age category over 10 simulations and hundreds of weeks after reaching the prevalence equilibrium and before vaccine introduction.

After calibration under the neutral scenario, we again calibrated the transmission-probability parameter $\beta_{NV}$ for each alternative interaction scenario, to reproduce the pre-vaccine NV-infection prevalence (Markowitz et al., 2013).

**Table S4.** Calibrated parameter descriptions, stratification, ranges of tested values and estimated values

| Symbol | Definition | Stratification | | Range | Estimated value |
|---|---|---|---|---|---|
| $P_{cg}$ | Percentage of individuals in each sexual activity level for $A_g = [15-17]$ <br> $c2 = 100 - c1 - c3$ | Men | c1 | [60–75] | 75 |
| | | | c3 | [3–10] | 6 |
| | | Women | c1 | [70–85] | 85 |
| | | | c3 | [3–6] | 3 |
| | Probability of partnership duration, % <br> c1 for $A_g = [15-17]$ | Men | –1 y | [25–60] | 50 |
| | | | [1–4] y | [5–25] | 15 |
| | | | [5–9] y | [5–25] | 15 |
| | | | 10+ y | [25–45] | 20 |
| | | Women | –1 y | [25–60] | 65 |
| | | | [1–4] y | [5–25] | 5 |
| | | | [5–9] y | [5–25] | 5 |
| | | | 10+ y | [25–45] | 25 |
| | Mean partnership or single-state duration | Men | *c2* | [8–16] | 14 |
| | | | *c3* | [2–6] | 5 |
| | | Women | *c2* | [8–16] | 14 |
| | | | *c3* | [2–6] | 3 |
| | Variance of partnership or single-state duration | *c2* | | | 8 (fixed) |
| | | *c3* | | | 5 (fixed) |
| $D_{mix}$ | Persistent partner-search duration before mixing between sexual activity levels (weeks) | *c*1 | | [5–20] | 10 |
| | | *c*2 | | | 5 |
| | | *c*3 | | | 30 |
| $MC_c$ | Maximum percentage of individuals who changed from c1 to another level and from c3 to another one | Men | | [30–98] | 45% |
| | | Women | | [30–98] | 95% |
| $M_{D\_im}$ | Mean immune-state duration | | | [8–14] | 12 |
| $\beta_{NV}$ | NV-type transmission-probability parameter | | | [0.05–3] | 0.125 |
| $\beta_V$ | V-type transmission-probability parameter | | | [0.05–3] | 0.16 |

### S3. Analysis of simulated results

#### S3.1 Partnership network

To describe our simulated partnership network, we defined the cumulative distributions of the total numbers of sexual partners of individuals from 18 to 30 years old. That cumulative distribution is given by the probability function that an individual had at least $x$ partners at a given time:

$$F_X(x) = P(X \geq x) = \sum_{x_i \geq x} P(X = x_i) = \sum_{x_i \geq x} p(x_i)$$

with $p(x_i)$ the percentage of individuals with $x_i$ partners. Cumulative distributions were calculated for both simulated and reported CSF data (Bajos and Bozon, 2008) and compared graphically.

#### S3.2 Infection pattern

To further characterise V- and NV-genotype distributions according to actual sexual activity group (0–1, 2–3 and >3 partners during the year), we computed the percentages of those not infected ("0"), those infected with a single genotype ("1") and those co-infected with more than one genotype (">1") in the whole population. Then, we restricted the analysis to the infected population to better understand how infected individuals are distributed across sexual activity groups. We computed the percentages of individuals infected with a single genotype, those co-infected with the two V genotypes ("V–V (NV = 0)"), with ≥2 NV genotypes ("NV–NV (V = 0)"), and those co-infected with both groups of genotypes ("V–NV").

Finally, to assess which behaviours were at transmission and/or acquisition risk, we computed the percentages of infections transmitted and acquired by individuals according to their sexual activity group.

##### S3.3 V- and NV-genotype prevalences

We defined V- and NV-genotype prevalences by age category $A$ as the ratio of the number of individuals infected with at least one V or NV genotype over the total number of individuals in the same age category.

$$P_A^V = \frac{N_A^V}{N_A} \qquad \& \qquad P_A^{NV} = \frac{N_A^{NV}}{N_A}$$

where $N_A^V$ and $N_A^{NV}$ are the numbers of individuals of age group $A$ infected with at least one V- and NV-type genotype, respectively, and $N_A$ is the number of individuals in that same age group $A$.

##### S3.4 Percentage differences: post- minus pre-vaccine prevalences

Here, we want to examine how a hypothetical interaction scenario would affect the ecology of HPV-genotype diffusion after vaccine introduction compared to the neutral scenario, in which no interaction occurs. To do so, we calculated the percentage differences, i.e., post- minus pre-vaccine prevalences of V and NV genotypes, for each interaction scenario and annual sexual activity group (0–1, 2–3 and >3 partners during the past year), as follows, respectively:

$$D^V = \frac{P_1^V - P_0^V}{P_0^V} \text{ and } D^{NV} = \frac{P_1^{NV} - P_0^{NV}}{P_0^{NV}}$$

With, respectively, $P_0^V$ and $P_0^{NV}$ the V- and NV-type prevalences before vaccine introduction and $P_1^V$ and $P_1^{NV}$ the V- and NV-type prevalences after vaccination was introduced for any given interaction scenario and real sexual activity group (0–1, 2–3 and >3 partners during the year). For each of the 20 simulations, pre- or post-vaccine, prevalence was calculated as the average of three time points each before and after vaccination was introduced after reaching prevalence equilibrium.

#### S4. Complementary results

##### S4.1 Partnership calibration: cumulative distributions of numbers of men's partners

The comparative distributions of total numbers of partners for men showed a small underestimation of the percentage of men with >3 partners (figure S3).

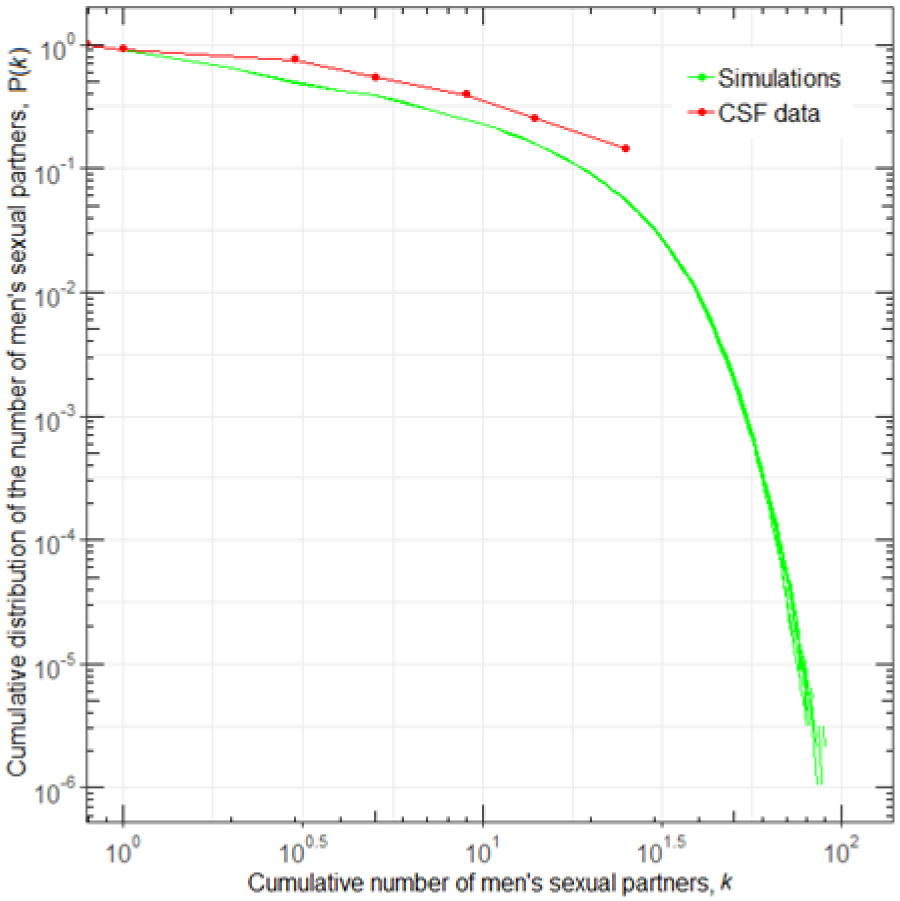

**Figure S3. Comparison of cumulative distributions of the numbers of sexual partners of men 18–30-years old in CSF data (Bajos and Bozon, 2008) and 10 model simulations.**

#### S4.2 Annual partnership-mixing matrix and comparison with virus-transmission matrix

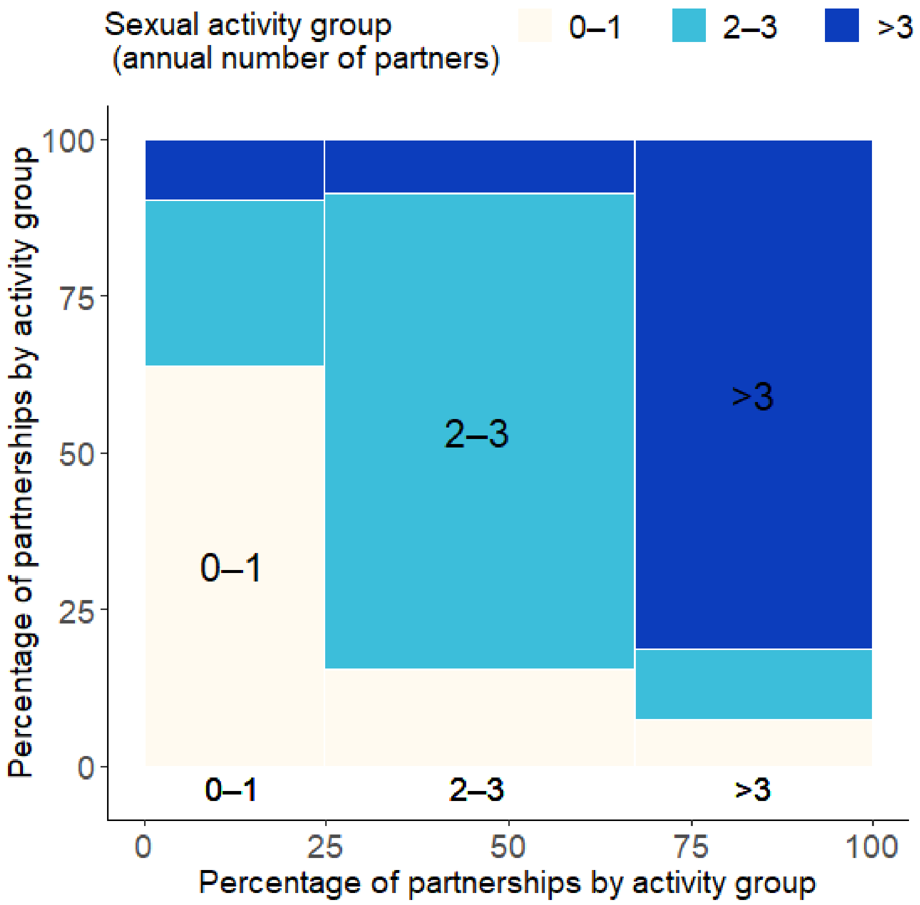

**Figure S4. Annual mixing matrix of partnership**. Percentages of new partnerships according to each partner's sexual activity group during the year.

We monitored individuals according to their number of partners during the past year: 0–1 partner (78.7% of the sexually active population representing 83.3% of the whole population), 2–3 partners (17.9%), >3 partners (3.4%). The annual mixing matrix shows that a large percentage of new partnerships (42.2% of individuals) are composed by individuals with 2–3 partners during the year (figure S4). The 3.4% of individuals having >3 partners were still involved in 33.2% of new couples. As expected, the percentage of new couples involving individuals with 0–1 partner was quite low. Notably, 75.0% of partnerships were formed with individuals from the same sexual activity group. Differences between transmission-acquisition patterns and annual mixing matrix of partnerships were observed for the lowest and highest sexually activity groups (figure 3C in main text and Figure S4). For individuals with 0–1 partner during the last year, the percentage of new couples more closely reflects to the percentage of viruses they acquired than the percentage of viruses they transmitted, which was lower (respectively, 24.6%, 22.9% and 12.9%). For individuals with 2–3 partners during the year, the percentage of new couples was comparable to those of transmitted and acquired viruses (respectively, 42.2%, 39.6% and 41.2%). Finally, for those who had >3 partners during the year, the percentage of new partnerships was closer to that of viruses they acquired than that of viruses they transmitted, which was higher (respectively, 33.2%, 35.9% and 47.5%).

##### S4.3 Calibration of genotype-interaction scenarios

For each interaction scenario, the probability of transmission was calibrated to reproduce realistic distributions of the prevalences of HPV-NV genotypes by age category, with the characteristic bell-shaped curve (V genotypes were previously calibrated for the neutral model; figure S5). Small variations in prevalence are observed across interaction scenarios.

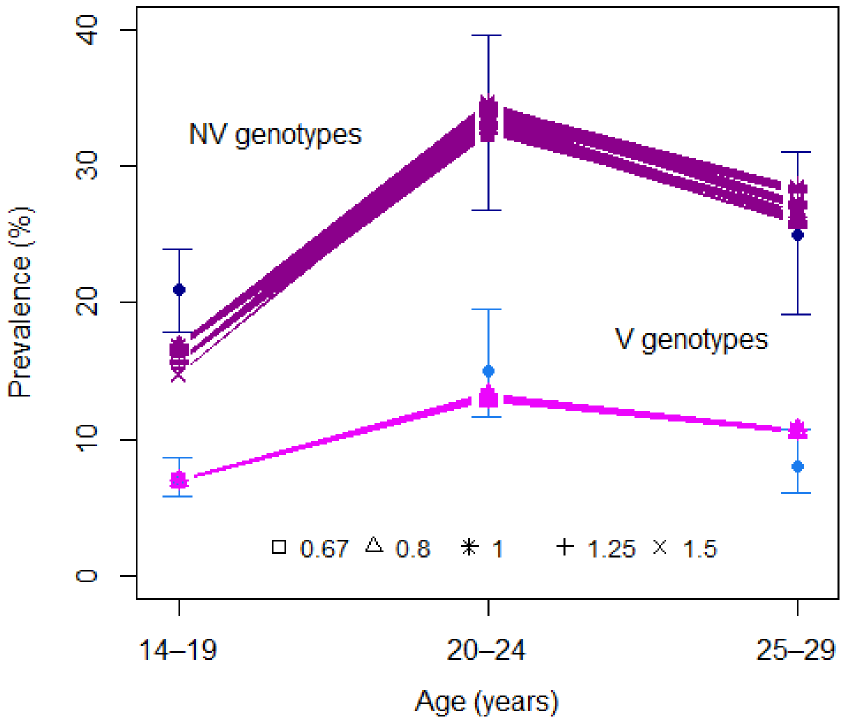

**Figure S5. Calibration of transmission parameters for each strength of interaction.** NV- and V-genotype prevalences for the observed data with 95% confidence interval *(Markowitz et al., 2013)* (dots and vertical bars) and for 10 simulation results for each strength of interaction (other symbols and connecting lines).

##### S4.4 Variations of NV- and V-genotype prevalences over time according to interaction scenario and sexual activity group

Figure 6 shows the evolution of V and NV prevalences over time before and after vaccine introduction. For each annual sexual activity group, V prevalences decreased similarly, regardless of the interaction scenarios. Conversely, NV-genotype prevalences changed markedly after vaccine introduction for all strengths of interaction compared to the neutral scenario, increasing for competitive scenarios and decreasing for synergistic scenarios. Nevertheless, NV-prevalence modifications were not observable for individuals with >3 partners during the year. New prevalence equilibria for V and NV genotypes was reached after 20 years.

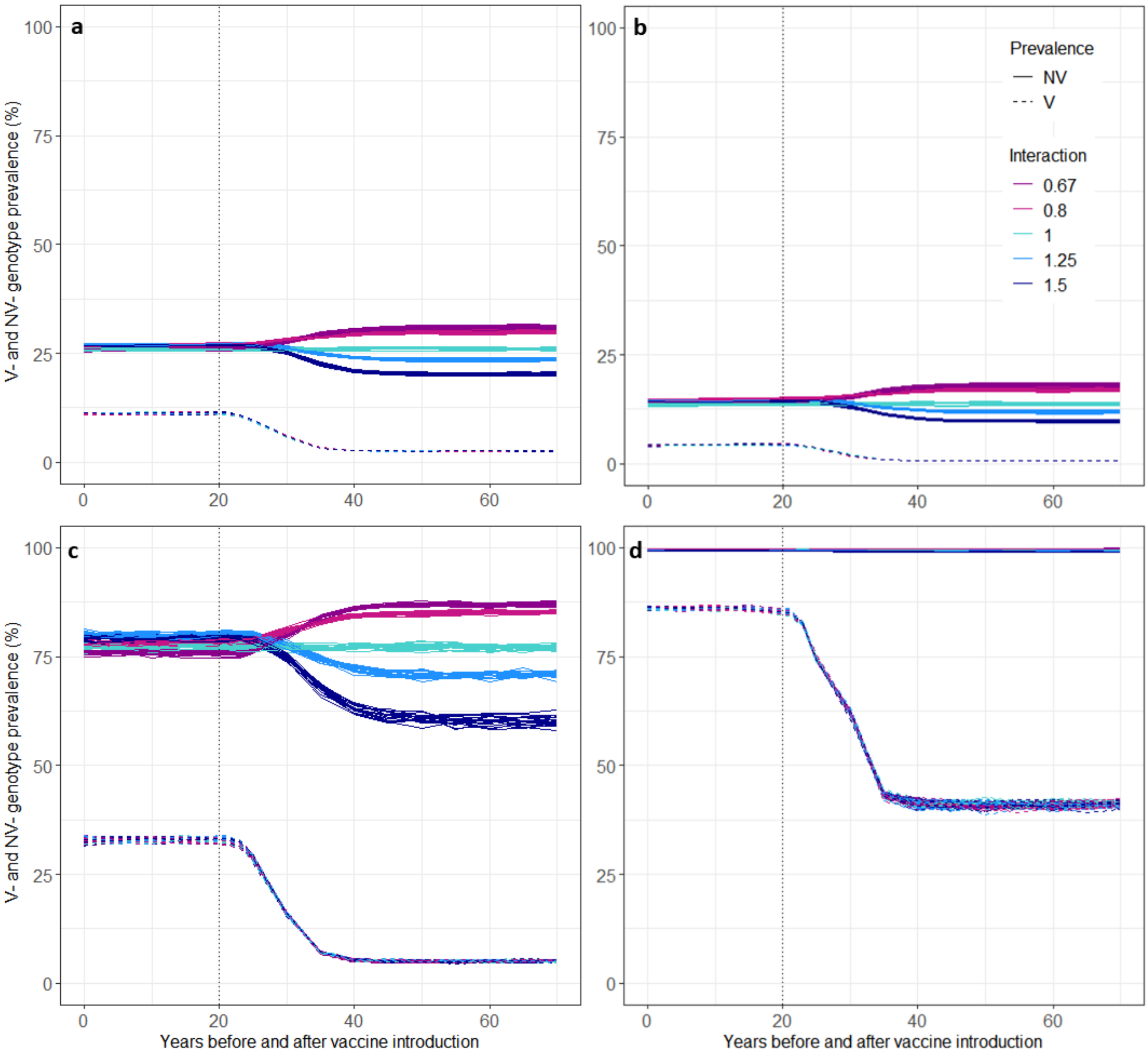

**Figure S6. NV- and V-genotype prevalences as a function of simulation time for each interaction scenario:** (a) All individuals; (b) Individuals with 0–1 partner during the past year; (c) 2–3 partners; (d) >3 partners. Vaccine was introduced at time *t* = 20 years ago for all women <15 years of age with 65% of vaccine coverage. For each interaction scenario, results of 20 simulations are displayed.

##### S4.5 Results for 25% uniform vaccine coverage for women

For V genotypes, the percentage differences between post-vaccine and pre-vaccine prevalences were the same, regardless of genotype interaction (figure 7), and they showed prevalences decreased by a median of 36.0% for all individuals. For competitive and synergistic interaction scenarios, respectively, the percentage differences between the pre- and post-vaccine prevalences indicated an increase or decrease of NV prevalence after vaccination. These modifications were stronger for greater interaction strengths and individuals with 0–1 partner (median 12.4% for $\alpha$ = 0.67 and –14.5% for $\alpha$ = 1.5) compared with individuals with 2–3 partners (median 8.2% for $\alpha$ = 0.67 and –9.5% for $\alpha$ = 1.5) and individuals with >3 partners (percentage differences <1%).

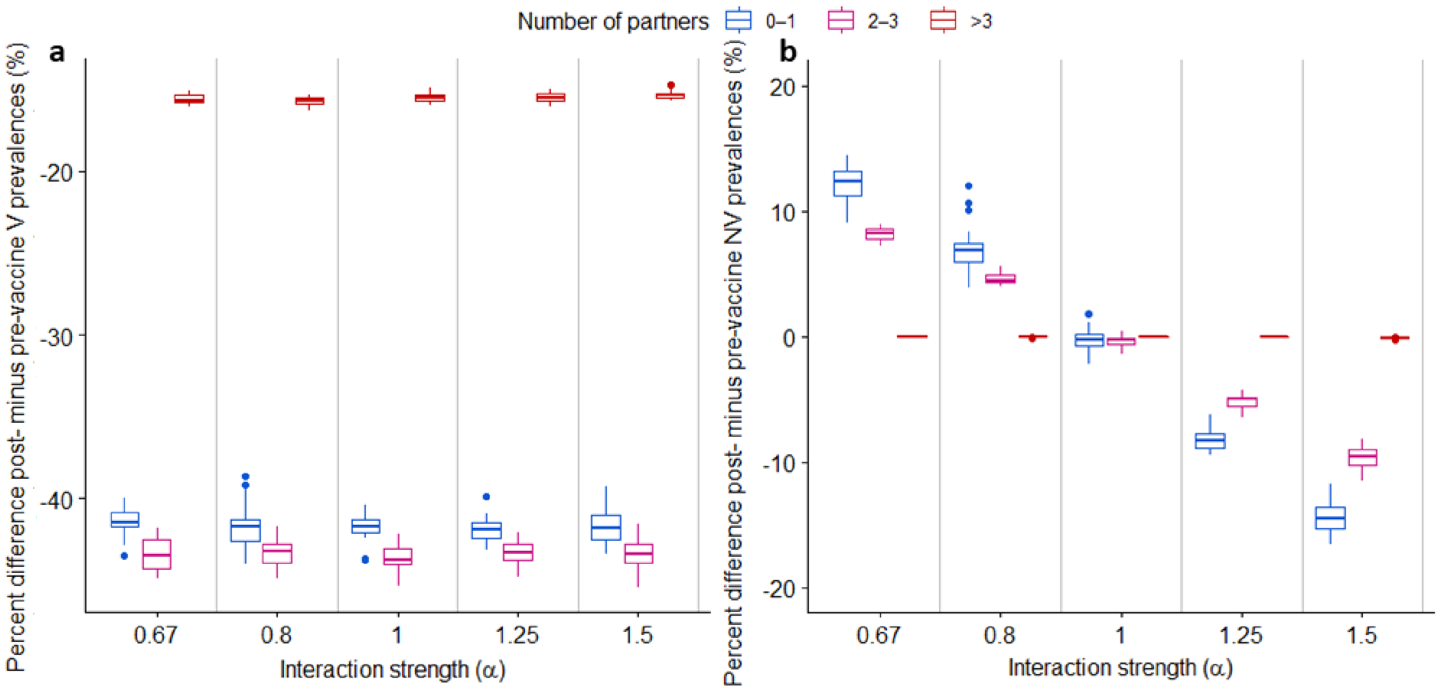

**Figure S7. Percentage differences of post- minus pre-vaccine (a) V- and (b) NV-genotype prevalences under 25% of vaccine coverage for all and by sexual activity group (0–1, 2–3, >3 partners during the year) and interaction scenarios (0.67, 0.8, 1, 1.25 and 1.5).** Vaccine was introduced at time *t* = 20 years ago. Average prevalences were assessed at three distinct times each before and after vaccine introduction. Boxplots display variability over 20 simulations for each interaction scenario (bold horizontal line inside the box is the median, lower and upper box limits are the 1st and 3rd quartiles, antennae correspond to 1.5 times the interquartile range and circles mark the values outside this range).

##### S4.6 Targeting the high-risk sexual activity group for vaccination

***Targeting the high-risk sexual activity group for vaccination did not modify NV-prevalence–variation patterns across sexual activity groups.*** We assessed the impact of targeting 15-year-old women with theoretically >3 partners for vaccination before a first partner, which represents ~3.4% of them. To better gauge the effect of this immunisation strategy, we compared it to a uniform vaccination scenario with same coverage. As expected, uniform vaccination of 3.4% of women had negligible impact on V- and NV-vaccine prevalence.

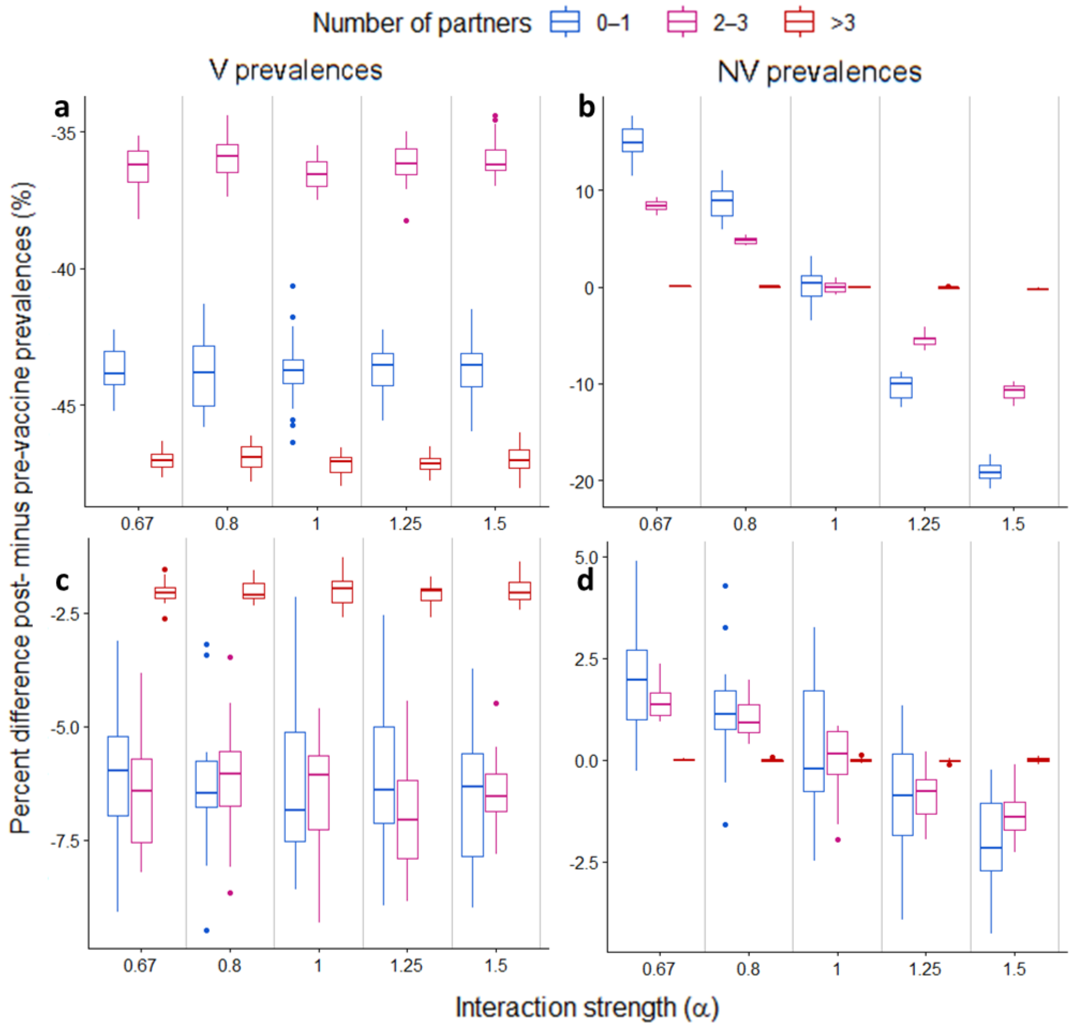

**Figure S8. Percentage difference between post- minus pre-vaccine (a & c) V- and (b & d) NV-genotype prevalences with (a & b) vaccination targeting 15-year-old women with >3 partners or (c & d) 3.4% uniform vaccine coverage for all and by sexual activity group and interaction scenarios (0.67, 0.8, 1, 1.25 and 1.5).** Vaccine was introduced at time *t* = 20 years ago. Average prevalences were assessed at three distinct times each before and after vaccine introduction. Boxplots display variability over 20 simulations for each interaction scenario (bold horizontal line inside the box is the median, lower and upper box limits are the 1st and 3rd quartiles, antennae correspond to 1.5 times the interquartile range and circles mark the values outside this range).

The percentage differences between post- and pre-vaccine prevalences were indeed close to 0 for all scenarios and sexual activity groups. Targeting highly active women led to a much stronger effect (figure S8a and c). Indeed, this vaccination strategy yielded a much lower V prevalence in individuals with >3 partners (median ratio: 0.53) than in those with fewer partners. Similarly, percentage differences between post- and pre-vaccine NV-genotype prevalences across all interaction scenarios were more marked under a targeted (figure S8b) as opposed to a uniform vaccination strategy (figure S8d). The patterns of NV-prevalence variation across sexual activity groups were similar to those observed under uniform immunisation strategy.

#### S5. Sensitivity analysis of the impacts of varying interaction strengths and transmission probability $\beta_{NV}$ on infection and co-infection patterns before and after vaccination

To assess the specific impact of genotype interaction on infection patterns without confounding by other parameter variations due to calibration, we simulated the model for all combinations of interaction strengths considered in the main analysis (0.67, 0.8, 1, 1.25 and 1.5) and transmission probability $\beta_{NV}$ obtained from the calibrated scenarios (0.072, 0.095, 0.125, O.21 and 0.28). Vaccination was introduced with 65% vaccine coverage after reaching prevalence equilibrium.

##### S5.1 Pre-vaccine infection and co-infection patterns

Before vaccine introduction (figure S9a–c), it can be seen that NV prevalence rose along with interaction strengths and the transmission probability $\beta_{NV}$ increases. In addition, the percentages of multiple NV infections and V–NV co-infections were similar for a given prevalence value obtained with different combinations of interaction strength and transmission probability $\beta_{NV}$. Thus, those findings confirmed what we observed for calibrated scenarios: i.e., that similar NV-prevalence and co-infection patterns can be obtained under synergistic, neutral and competitive interaction scenarios.

##### S5.2 Post-vaccine infection and co-infection patterns

***Only NV–V co-infection among NV-infected individual distinguished competitive from synergistic scenarios.*** Comparing the figures before and after vaccine introduction, we can observe that NV prevalence decreased or increased, respectively, in the synergistic or competitive interaction scenario (figure S9). Nevertheless, the same prevalence level could still be obtained for different interaction strengths. Moreover, the post-vaccine percentage of NV co-infections among individuals infected with NV genotypes kept the same proportionality coefficient as during the pre-vaccine era. Following vaccine introduction, only the percentage of NV–V co-infections was impacted differently depending on whether the interactions were competitive or synergistic. The decrease of V–NV co-infections in percentage to all NV infections was more pronounced in competitive than synergistic scenarios

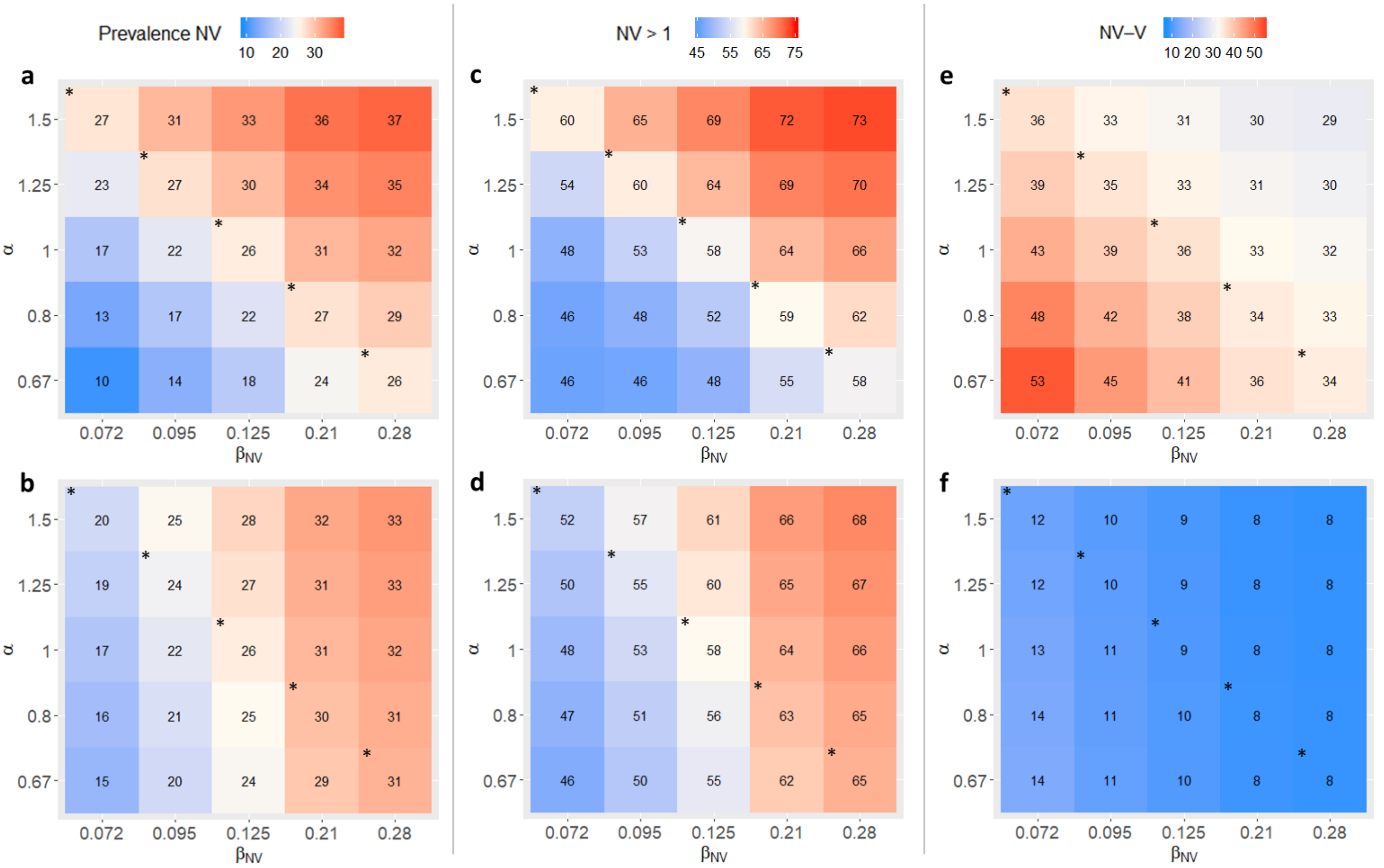

**Figure S9. NV-genotype prevalences and percentages of NV, NV > 1 and NV–V genotype co-infections among those infected with NV before and after vaccination, according to interaction-strength values ($\alpha$) and transmission probability $\beta_{NV}$.** Mean NV prevalences over 20 simulations (a and b); among NV-infected individuals, percentages (%) of individuals infected with >1 NV genotypes (c and d) or NV–V co-infected (e and f), before (a, c and e) and after vaccination (b, d and f). Average percentages (rounded values in each box) were assessed at three distinct times each before and after vaccine introduction and 20 simulations for each interaction-strength combination and transmission probability $\beta_{NV}$. (*for calibrated scenarios).

#### S5.3 Percentage differences: NV post- minus pre-vaccine prevalences

Considering percentage differences between NV post- and pre-vaccine prevalences, we confirm that competition or synergy scenarios, respectively, led to higher or lower NV prevalences compared with the neutral model (Figure. S10 a). For a given transmission probability $\beta_{NV}$, the stronger the interaction strength (the more it deviates from 1), the greater the percentage difference of NV prevalences. Interestingly, we found a positive correlation between the absolute percentage differences between post- and pre-vaccine NV prevalences and the percentage of NV-V coinfections among NV infected individuals before vaccination (figure S10b, Spearman's rank correlation coefficient $r$ = 0.84).

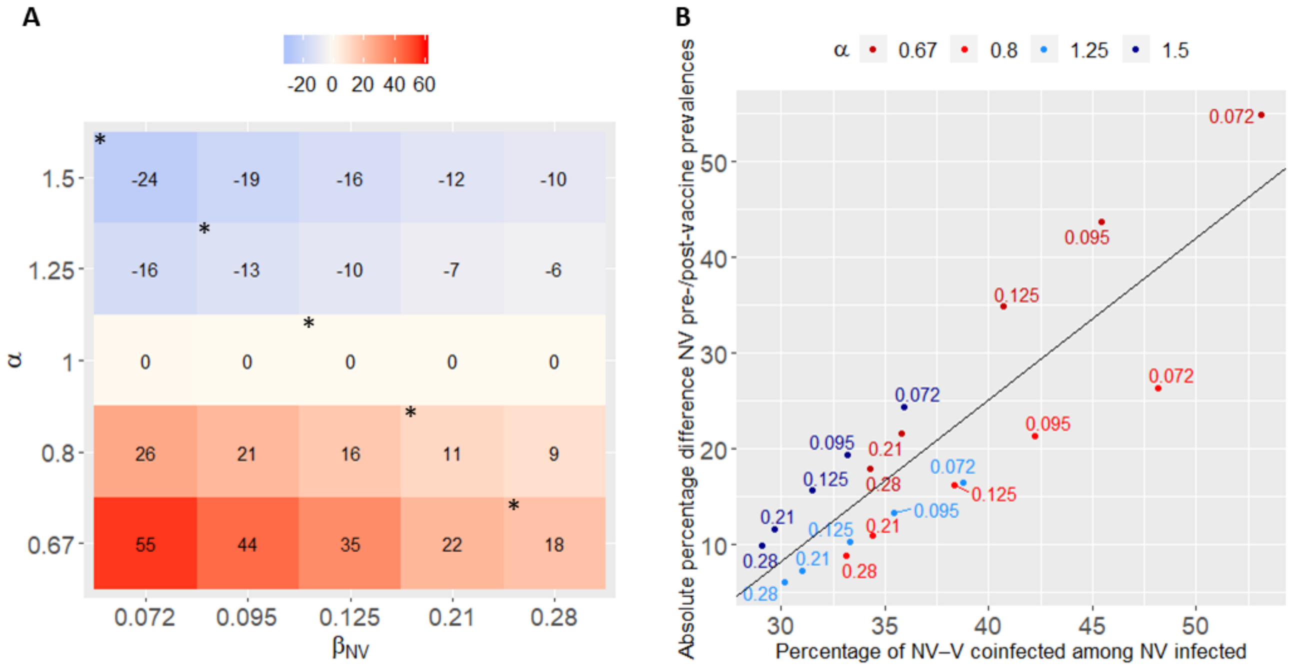

**Figure S10. NV-genotype post- minus pre-vaccine prevalences and NV–V co-infection.** (a) The mean percentage difference between NV-genotype post- and pre-vaccine prevalences, according interaction strength ($\alpha$) and transmission probability $\beta_{NV}$. Vaccine was introduced at $t$ = 20 years ago. Mean prevalences were assessed by averaging three distinct times each before and after vaccine introduction over 20 simulations for each scenario (*for calibrated scenarios). (b) The absolute NV-prevalence differences between pre- and post-vaccine according to the percentage of NV–V co-infection among NV-infected individuals before vaccination. Circles are simulated values for interaction-strength (in red for competitive interaction and blue for synergistic interaction) and transmission probability $\beta_{NV}$ combinations (values written next to the circle. The straight line is to the regression line of the linear model associated with observations of the form $y$ = 1.687x – 42.317. The Spearman correlation coefficient was 0.84 with method ($p = 2.2\times10^{6}$).

### S6. Sensitivity analysis of between-genotype interactions

#### S6.1 Definitions of interaction scenarios

In the sensitivity analysis, we considered two mechanisms of interaction, whereby ongoing infection by a genotype changes either the probability of acquisition of another genotype or the duration of infection with another genotype by a multiplicative factor (table S5). When the interaction mechanism affected the probability of acquiring the genotype $P_{Acq}$ (mechanism called "Acq"), the probability was defined such that $P_{Acq}^{eff} = P_{Acq} \times \alpha_P$ with $\alpha_P$ the interaction parameter with respect to acquisition probability. When the interaction mechanism affected type-specific duration of infection $D_{INF}$ (mechanism called "Dur"), the duration was defined such that $D_{INF}^{eff} = D_{INF} \times \alpha_D$ with $\alpha_D$ the interaction parameter with respect to duration. Three degrees of between-type interactions were embedded:

- Unilateral interaction, whereby V-genotype infection changes consecutive infection with NV type
- Bilateral interaction, whereby NV-genotype infection may change consecutive infection with V type and vice versa
- Universal interaction, whereby any genotype infection may change consecutive infection with any other type of the same or the other V/NV group.

**Table S5.** Definition of scenarios of interaction that can alter either the duration of infection or the probability to acquire following three degrees of between-types interaction

| Interaction mechanisms | Three degrees of between-type interactions | Ecological interaction & scenario label | |
|---|---|---|---|
| | | Competitive (C) | Synergistic (S) |
| Changes infection duration (Dur) | 1) NV-genotype acquisition in the presence of a V genotype | 1C_Dur | 1S_Dur |
| | 2) acquisition of a genotype of one group in the presence of a genotype of the other group | 2C_Dur | 2S_Dur |
| | 3) acquisition of a genotype in the presence of at least one other genotype | 3C_Dur | 3S_Dur |
| Changes the probability to acquire (Acq) | 1) transmission of an NV genotype in the presence of a V genotype | 1C_Acq | 1S_Acq |
| | 2) transmission of a genotype of one group in the presence of a genotype of the other group | 2C_Acq | 2S_Acq |
| | 3) transmission of a genotype in the presence of at least one other genotype | 3C_Acq | 3S_Acq |
| | | Neutral N | |

#### S6.2 Calibration

After calibrating the neutral scenario, alternative scenarios, in which the interaction concerned infection duration, were first optimised for a fixed interaction parameter $\alpha_D$ at 0.8 for competition or 1.25 for synergy scenario by calibrating transmission probabilities $\beta_{NV}$ and $\beta_V$ (only for bilateral and universal interactions). To ensure similar interaction levels, in scenarios in which interaction concerned the probability of acquisition, the transmission probabilities were fixed at the values obtained from the comparable interaction scenario of infection duration (competitive or synergistic, unilateral, bilateral or universal), while the interaction strength $\alpha_P$ was optimised to reproduce V and NV prevalences. Calibration results are presented in table S6. The model could be satisfactorily calibrated against real data for all scenarios except 3S_Dur and 2C_Acq, as reflected by greater least-squares distances than in other interaction scenarios.

**Table S6.** Calibration of interaction scenarios

| Interaction name | Interaction value[a] | $\beta_{NV}$ [a] | $\beta_{V}$[a] | Sum of least-squares distance minimized* | Least-squares distance minimized for NV* | least squares distance minimized for V* |
|---|---|---|---|---|---|---|
| N | 1 | **0.125** | **0.16** | 0.0038 | 0.0028 | 0.0010 |
| 1C_Dur | 0.80 | **0.21** | 0.16 | 0.0037 | 0.0027 | 0.0010 |
| [b] | 0.67 | **0.28** | 0.16 | 0.0031 | 0.0020 | 0.0011 |
| 1S_Dur | 1.25 | **0.095** | 0.16 | 0.0050 | 0.0039 | 0.0011 |
| [b] | 1.50 | **0.072** | 0.16 | 0.0062 | 0.0051 | 0.0011 |
| 2C_Dur | 0.80 | **0.20** | **0.80** | 0.0033 | 0.0019 | 0.0014 |
| 2S_Dur | 1.25 | **0.096** | **0.075** | 0.0051 | 0.0035 | 0.0016 |
| 3C_Dur | 0.80 | **0.70** | **1.00** | 0.0031 | 0.0017 | 0.0014 |
| 3S_Dur | 1.25 | **0.07** | **0.075** | 0.0071 | 0.0054 | 0.0017 |
| 1C_Acq | **0.36** | 0.21 | 0.16 | 0.0039 | 0.0028 | 0.0011 |
| 1S Acq | **2.80** | 0.095 | 0.16 | 0.0040 | 0.0029 | 0.0011 |
| 2C_Acq | **0.425** | 0.20 | 0.80 | 0.0076 | 0.0031 | 0.0045 |
| 2S Acq | **2.50** | 0.096 | 0.075 | 0.0040 | 0.0030 | 0.0010 |
| 3C_Acq | **0.15** | 0.70 | 1.00 | 0.0038 | 0.0027 | 0.0011 |
| 3S Acq | **2.50** | 0.07 | 0.075 | 0.0043 | 0.0033 | 0.0010 |

*Optimised value is the sum of least squares between data and simulation results. [a] In bold are the fitted value(s) for each model. [b] Interaction scenarios of infection duration considered in the main analysis but not integrated into the sensitivity analysis.

#### S6.3 Comparison of interaction scenarios before and after vaccination

After vaccination introduction, the assumptions made regarding genotype interactions strongly impacted the V- and NV-prevalence trends (figures S11 and S12). Results are presented here for 25% and 65% vaccine coverages.

For the competition and synergistic scenarios, the percentage differences between post- and pre-vaccine V prevalences showed their decreases to be similar to those under the neutral scenario, except when interaction pertained to infection duration in a bilateral or universal manner (2C_Acq, 2C_Dur, 2S_Dur, 3C_Dur and 3S_Dur). In those cases, we observed a stronger reduction for synergistic interactions and a weaker reduction for competitive scenarios. Differences were more pronounced with 65% than 25% vaccine coverage. Before vaccine introduction, we observed that, under scenario 2C_Acq, V prevalence was higher (median 17.5) than under the neutral interaction scenario (median 13.6, figure S13a). In contrast, V prevalence was lower under 2C_Dur, 2S_Dur, 3C_Dur and 3S_Dur scenarios (median range 12.0–13.0). These V-prevalence differences with the neutral scenario could explain the outcomes observed after vaccine introduction.

Interestingly, changes post-vaccine introduction differed according to sexual behavior characteristics. They were more pronounced for individuals with ≤3 partners than the others. Thus, for individual with ≤3 partners, the median prevalence-difference percentage declined by 84.6% of the post-vaccination prevalence for 65% of vaccine coverage (42.5%), while for individuals with >3 partners, the decrease was around 52.3% for 25% of vaccine coverage (15.5%).

The percentage NV-genotype–prevalence differences, i.e., post- minus pre-vaccine changed significantly between the two periods for all scenarios, except the neutral interaction and for individuals with >3 partners during the year (percent modification <1%). The percentage of NV-prevalence differences differed significantly from 0 only for 65% vaccine coverage, when interactions were assumed to occur between all genotypes (3C_Dur, 3C_Acq, 3S_Dur and 3C_Acq, <5%). The latter result could be expected because prevalence decreased for only two V genotypes (compared to 12 NV genotypes) following vaccination, so co-infections were not significantly diminished, and made no notable impact on interactions among all genotypes within the considered range of interaction strengths. Generally, the prevalence-difference percentages were more pronounced with heightened vaccine coverage and fewer partners.

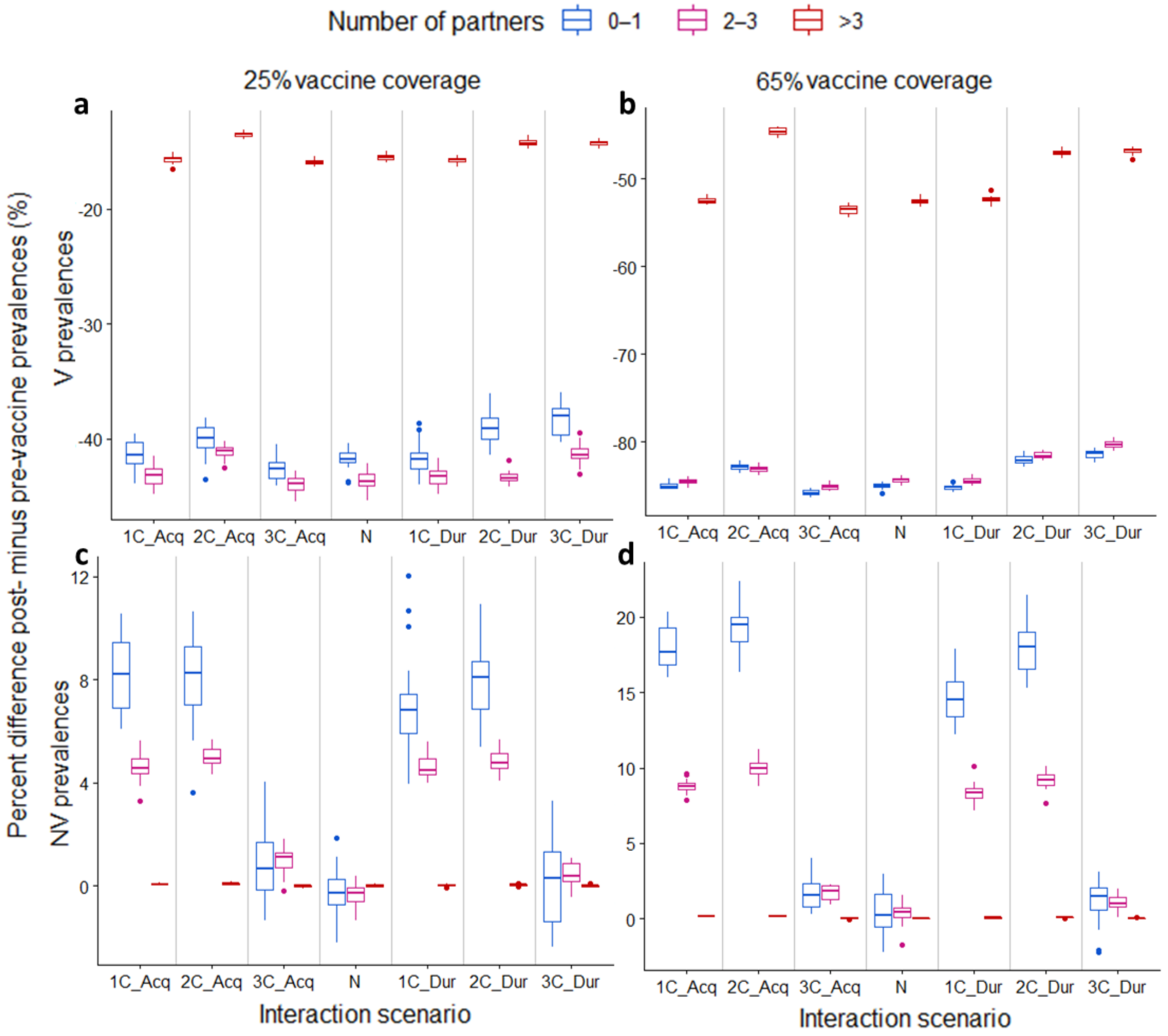

**Fig S11. Percentage differences: post- minus pre-vaccine (a and b) V- and (c and d) NV-genotype prevalences for competitive and neutral interaction scenarios under 25% (a and c) and 65% (b and d) vaccine coverage by group of annual number of partners (0–1, 2–3, >3) during the year.** Vaccine was introduced at time t = 20 years ago. Average prevalences were assessed at three distinct times each before and after vaccine introduction. Boxplots report variability over 20 simulations for each interaction scenario (bold horizontal line inside the box is the median, lower and upper box limits are the 1st and 3rd quartiles, antennae correspond to 1.5× the interquartile range and circles mark the values outside this range).

For competitive interaction scenarios (figure S11), the percentages of NV-prevalence differences indicated prevalence increases after vaccination, stronger for individuals with 0–1 partners with 65% or 25% coverage, respectively: median ranges 14.6%–19.5% and 6.8%–8.2%, than for individuals with 2–3 partners: median range 8.4%–10.0% and 4.5%–4.9%. In contrast, the percentages of NV-prevalence differences for synergistic scenarios indicated decreased prevalence after vaccine introduction (figure S12). Again, for respective 65% or 25% coverage, the extent of decreases was more pronounced for individuals with 0–1 partner (median range between –18.2% to –15.0% and –8.3 to –6.0%) than individuals with 2–3 partners (median ranged between –12.0% and –9.0 %: –5.0 and –3.9).

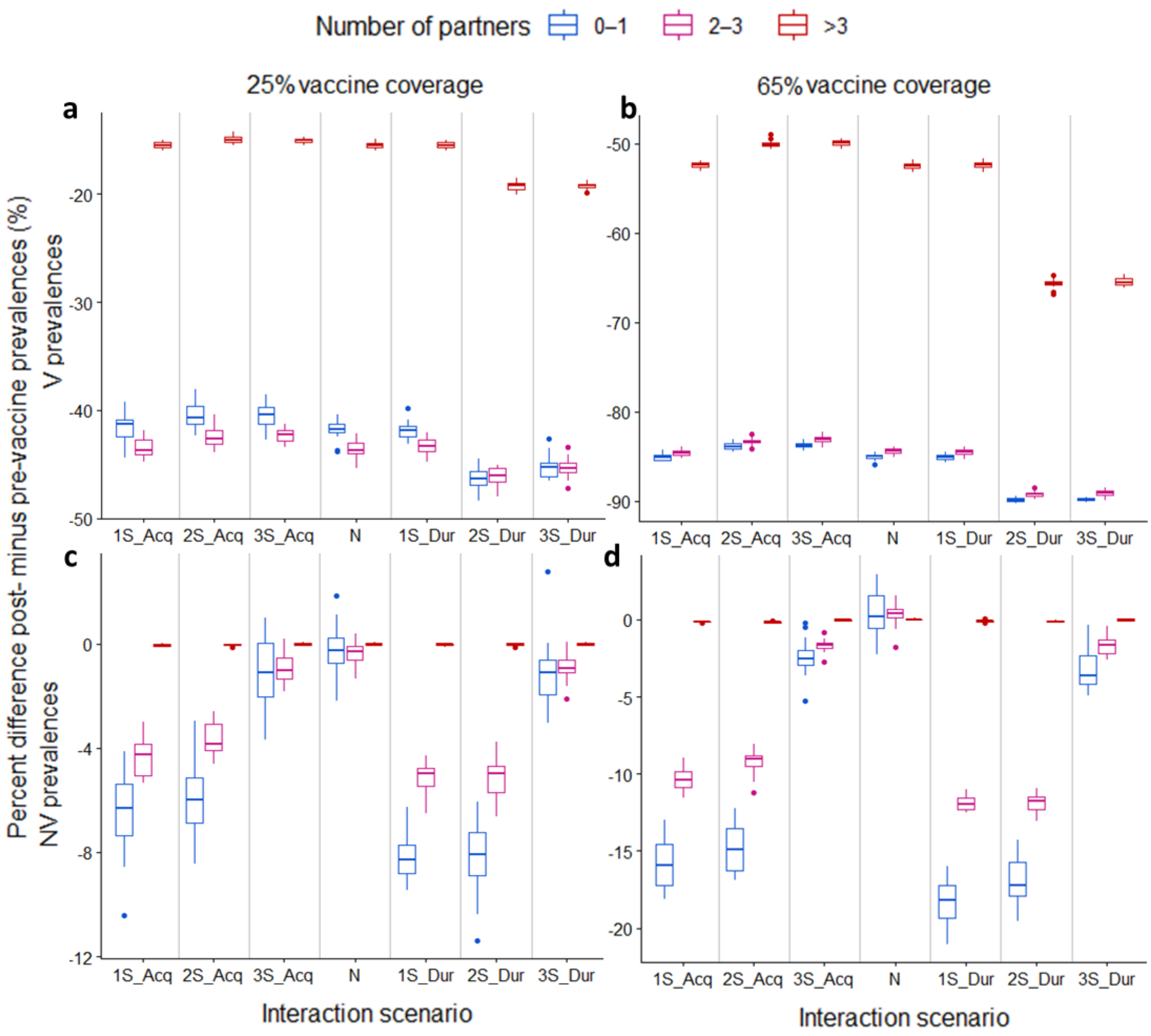

**Figure S12. Percentage differences: post- minus pre-vaccine (a and b) V- and (c and d) NV-genotype prevalences for synergistic and neutral interaction scenarios under 25% (a and c) and 65% (b and d) vaccine coverage by group of annual number of partners (0–1, 2–3, >3) during the year.** Vaccine was introduced at t = 20 years ago. Average prevalences were assessed at three distinct times each before and after vaccine introduction. Boxplots show variability over 20 simulations for each interaction scenario (bold horizontal line inside the box is the median, lower and upper box limits are the 1st and 3rd quartiles, antennae correspond to 1.5× the interquartile range and circles mark the values outside this range).

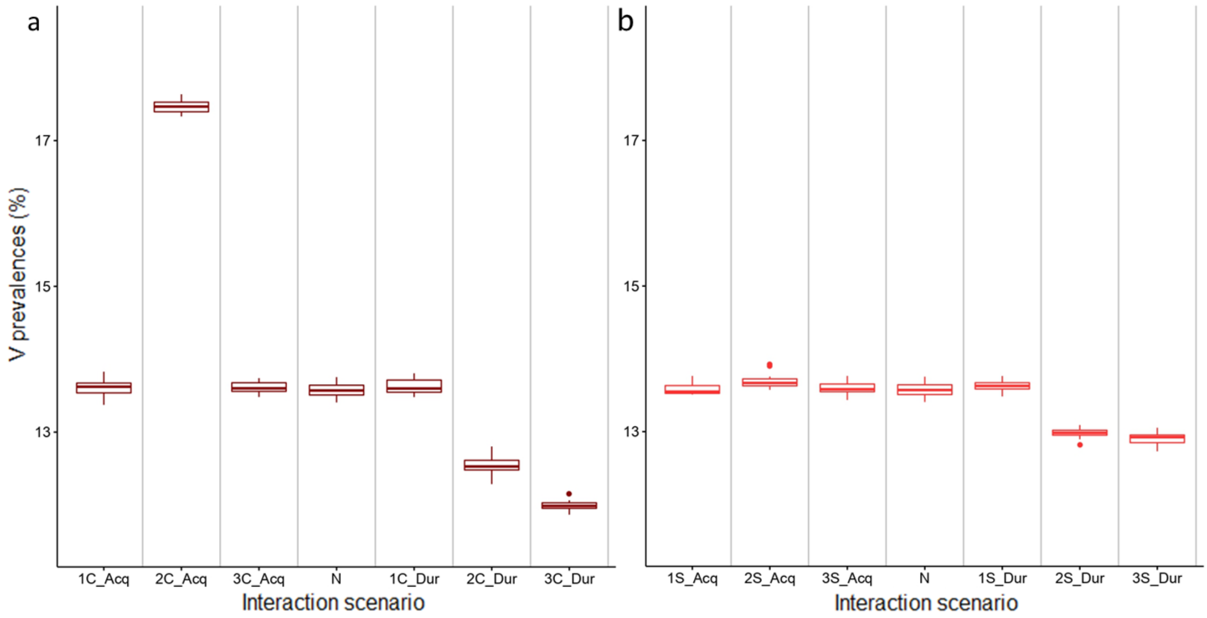

**Figure S13. Percentage V-genotype prevalences for the neutral, competitive (a) and synergistic (b) scenarios among the sexually active population**. Average prevalences were assessed at three distinct times before vaccine introduction. Boxplots report variability over 20 simulations for each interaction scenario (bold horizontal line inside the box is the median, lower and upper box limits are the 1st and 3rd quartiles, antennae correspond to 1.5× the interquartile range and circles mark the values outside this range).
